# Impact of the representation of contact data on the evaluation of interventions in infectious diseases simulations

**DOI:** 10.1101/2022.02.28.22271600

**Authors:** Diego Andrés Contreras, Elisabetta Colosi, Giulia Bassignana, Vittoria Colizza, Alain Barrat

## Abstract

Computational models offer a unique setting to test strategies to mitigate infectious diseases’ spread, providing useful insights to applied public health. To be actionable, models need to be informed by data, which can be available at different levels of detail. While high resolution data describing contacts between individuals are increasingly available, data gathering remains challenging, especially during a health emergency: many models thus use synthetic data or coarse information to evaluate intervention protocols. Here, we evaluate how the representation of contact data might affect the impact of various strategies in models, in the realm of COVID-19 transmission in educational and work contexts. Starting from high resolution contact data, we use data representations ranging from very detailed to very coarse to inform a model for the spread of SARS-CoV-2 and simulate several mitigation strategies. We find that coarse data representations underestimate the risk of super-spreading events. However, the rankings of protocols according to their efficiency or cost remain coherent across representations, ensuring the consistency of model findings to inform public health advice. Caution should be taken, however, on the quantitative estimations of those benefits and costs that may trigger the adoption of protocols, as these may depend on data representation.

## 1 Introduction

Computational models and numerical simulations are essential tools for the understanding of epidemic spread [1, 2], at scales ranging from global to local [3, 4, 5, 6]. They have been used in the past to examine pandemic scenarios, and more extensively during the current COVID-19 pandemic, to evaluate the potential impact of non-pharmaceutical interventions (NPIs) ranging from international travel restrictions [5, 4, 7, 8, 9] to lockdowns or curfews aiming at reducing global mobility and interactions [10, 11, 12, 13], to more targeted measures such as isolation of positive cases, contact tracing, telework, partial closures of schools or surveillance by regular testing [14, 15, 16, 17, 18, 19, 20, 21, 22].

Epidemic models of infectious diseases rely on the one hand on the physiological characterization of the disease within hosts, and on the other hand on the representation of how the disease can propagate from host to host, i.e., of the interactions between hosts. These interactions can be described at various levels of details: at the coarsest level, homogeneous mixing [1] assumes that all individuals can interact with each other in a uniform way; contact matrices divide individuals into classes, and give the average duration of contacts between individuals of given classes [23]; contact networks describe specifically which pairs of hosts have been in contact [24, 25, 26]. Regardless of the level of description chosen, a model needs to be informed by data in order to be actionable, i.e., to provide scenarios that can inform public health decisions. These data are typically collected by surveys or diaries [27, 23, 28] or, more recently, using wearable sensors able to detect close-range proximity between individuals with high spatial and temporal resolution [29, 30, 31, 32, 33].

Gathering data is however expensive, time-consuming and implies logistical challenges, which become particularly prohibitive for large-scale populations or multiple coupled settings, especially for high-resolution data [25, 34]. The questions of how much detail should be included in computational models, and of how the description of interactions impacts the outcome and actionability of computational models, arise therefore naturally [6, 35]. For instance, the estimation of the role of superspreading needs to be informed by the heterogeneity of contact patterns [36]. Coarse representations can also overestimate the impact of a spread or the epidemic risk of specific groups [6, 37, 38], even if a rescaling of parameters can enhance the accuracy of models based on a homogeneous mixing hypothesis [39]. To overcome the limitations of coarse representations, intermediate data representations informed by statistical heterogeneities of contact numbers and duration yielding a good estimation of the epidemic risk have been put forward [37, 38].

Although data with a limited resolution were shown to be insufficient to inform interventions at individual scale [40], they are still useful to inform strategies at intermediate scales [41, 42, 14, 15, 43]. In practice however, a general issue faced by models concerns the comparison of strategies or control measures, in terms of both costs and benefits. In the case of COVID-19 for instance, the computational models mentioned above have considered a wide variety of measures (contact tracing, regular testing, telework, class or school closures), with each study using specific empirical or synthetic data and a specific representation of contacts [17, 20, 44, 45, 22, 19, 46, 47, 48, 21, 43]. However, just as the data representation can affect the identification of risk groups [37], it might also impact the ranking of different strategies. Here we tackle this issue by leveraging high-resolution data describing contacts between individuals in several settings (offices, schools, hospital). We consider several representations of the data, from fine-detailed to coarse-grained [37], and use these representations to feed an agent-based model of propagation of SARS-CoV-2 infection in these settings. We simulate several strategies (reactive and regular testing, telework, reactive class closures) and evaluate their cost and benefit for each representation, highlighting differences and similarities in the outcomes.

## 2 Methods

We consider a model for the spread of COVID-19 in different settings, namely two distinct school settings, an office setting and a hospital ward. In this section, we first present the compartmental model used and the non-pharmaceutical interventions (NPI) considered. We then describe the high-resolution data on interactions between individuals that we use, as well as the hierarchy of coarse-grained representations of the contact patterns that preserve the temporal and structural information of the data at different levels of detail.

### 2.1 Compartmental model

We use an agent-based model in which the progression of the disease within each host follows discrete states, as shown in Figure 1a [20]. Infectious individuals can transmit the disease to susceptible (healthy) individuals (*S*), who first enter the exposed (non-infectious) state (*E*) and then a pre-symptomatic infectious state (*I*_*p*_) after a time *τ*_*E*_. The pre-symptomatic phase lasts *τ*_*p*_, after which individuals either evolve into a sub-clinical infection (*I*_*sc*_) or manifest a clinical infection *I*_*c*_, with respective probabilities 1 − *p*_*c*_ and *p*_*c*_. The infectious state leads finally to the recovered state *R* after a time *τ*_*I*_. The disease state durations *τ*_*E*_, *τ*_*p*_ and *τ*_*I*_ are distributed according to Gamma distributions, with average values and standard deviations given in Table 1 (See also Supplementary Material - SM, Section S1.2.4).

**Table 1:** Parameters of the compartmental model, taken from [20]

| SEIR parameter | value |
| --- | --- |
|  | mean (std) [days] |
| $\tau_E$ | 4 (2.3) |
| $\tau_P$ | 1.8 (1.8) |
| $\tau_I$ | 5 (2.0) |
| $R_0$ | 1.5, 3.0 |
| $p_c$ | 0.5 |
| $\sigma$ | 1.0 |
| $r_\beta^p, r_\beta^{sc}$ | 0.55 |
| $r_\beta^c$ | 1.0 |

**Figure 1:**
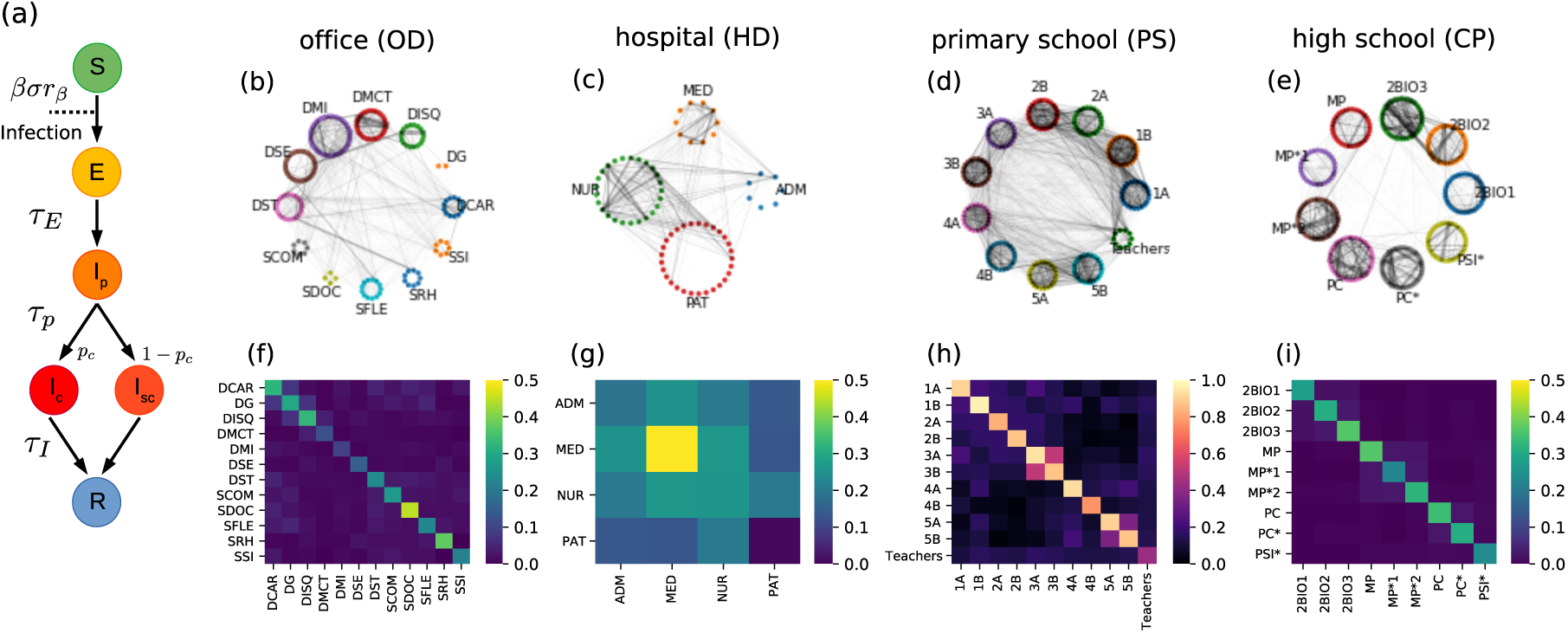
Model and data sets. (a) Schematic illustration of the epidemic model. After contact with an infected individual, a susceptible individual can become exposed, then transition to a pre-symptomatic state, which develops into either a clinical or a sub-clinical infection before recovering. (b,c,d,e) Weighted networks of contacts for each setting, aggregating in each case the interactions over the first day of the data collection in the settings OD, HD, PS and CP, respectively. The width of an edge is proportional to its weight, i.e., the total contact time between the individuals connected. (f,g,h,i) Contact matrices showing the average daily density of links between categories in the settings OD, HD, PS and CP, respectively. Namely, if there are 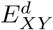 links on day *d* between the *N*_*X*_ individuals of category *X* and the *N*_*Y*_ individuals of category *Y*, the (*X, Y*) element of the contact matrix is given by the average over days of 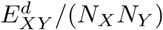, (if 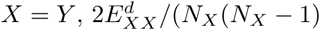).

Transmission of the disease can occur upon contact between a susceptible and an infectious (*I*_*p*_, *I*_*sc*_ or *I*_*c*_). The probability of transmission per unit of time depends on the product of the transmission rate *β*, the relative infectiousness *r*_*β*_ of the infectious individual and the susceptibility *σ* of the agent. The parameter *β* is tuned to obtain a desired specific value for the basic reproductive number *R*_0_, as detailed in the SM Section S1.3. The relative infectiousness *r*_*β*_ depends on the compartment of the infectious individual, with a larger 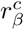 value for infectious individuals in the clinical state *I*_*c*_, and lower values 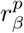 and 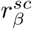 for *I*_*p*_ and *I*_*sc*_ (Table 1). It also depends on the age class of the infectious, with adults and adolescents more infectious than children (Table 2). The susceptibility *σ* also depends on the age of the susceptible individual, with adults more susceptible than other groups (adolescents and children have a susceptibility reduced by respectively 25% and 50% with respect to adults, see Table 2). Finally, the probability to develop a clinical infection is also reduced by 60% for both adolescents and children.

**Table 2:** Reduction in susceptibility *σ*, probability of clinical infection *p*_*c*_ and relative infectiousness *r*_*β*_ for children and adolescents, with respect to their values for adults. Taken from [20]

| parameter | reduction for<br>children | reduction for<br>adolescents |
| --- | --- | --- |
| $\sigma$ | 50 % | 25% |
| $p_c$ | 60 % | 60% |
| $r_\beta$ | 27 % | 0 % |

We can further enrich the compartmental model of Figure 1a by considering that individuals can be vaccinated. Here we do not consider a dynamic vaccination rollout, and assume that vaccination coverage is fixed throughout the simulation. We also assume full vaccination of individuals. We assume vaccination to reduce *r*_*β*_ by 50%, *σ* by 85%, and *p*_*c*_ by 93% We consider (in the SM, Section S2.4) levels of vaccination coverage of 25%, 50%, and 75%. As sensitivity analysis, we also consider a less effective vaccine (see SM Section S2.5.3).

### 2.2 Non-pharmaceutical interventions

We consider several interventions based on testing and isolation of cases, as well as closure of classes in school settings, and telework in offices. We first use as baselines the following cases:

- **No Test (NT):** No control strategies are in place. This would correspond to uncontrolled disease spread.
- **Symptomatic testing (ST) and case isolation:** Clinical cases have a probability *p*_*D*_ = 0.5 (*p*_*D*_ = 0.3 for children) to take a test and then isolate for Δ_*Q*_ = 7 days after receiving the result of the test. Tests are performed outside work/school hours. Symptomatic individuals remain isolated while they wait for their test results.

The NT case is used for calibration purposes and to study general properties of an unrestrained spread. The ST protocol is used as a reference protocol against which all other protocols are compared. With ST always implemented, we consider the following additional NPIs:

- **Regular testing (ST + RT** *α t/w*): Non-vaccinated individuals are periodically tested *t* times every *w* weeks with an adherence *α*. Positive cases remain in isolation for Δ_*Q*_ = 7 days. Tests are performed during work/school hours.
- **Telework (ST + TW** *d/w***) :** Telework is implemented in rotation on groups of individuals, only in the office setting. Each group works remotely once every *g* working days. Each individual stays thus at home *d* = 5*/g* times a week, and each working day a fraction (*g* − 1)*/g* of individuals is present at the office. We consider, for simplicity, only *g* = 5 and *g* = 10, corresponding to one day of telework every week or every two weeks.
- **Class quarantine (ST + Qc):** This protocol is implemented only in the school settings. When an individual is tested positive through the ST intervention, the whole class goes into isolation for Δ_*Q*_ = 7 days.
- **Reactive testing (ST + rT** *α*): This protocol is implemented only in the school settings and in the office setting. When an individual tests positive through the ST intervention, the non-vaccinated students of the same class (for schools) or the members of the same department (for offices) are tested after a time Δ_*r*_ = 1 day, with an adherence *α*. A second test is performed after 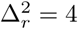 days. Positive cases are quarantined during Δ_*Q*_ = 7 days.

In the office setting, we additionally consider a protocol in which RT is combined with TW, (RT+TW). Further details of the implementation can be found in the SM Section S1.2.

The efficacy of a protocol is quantified in terms of relative reduction of cases with respect to the ST at the end of 60 simulation days. The cost is measured as the average number of days spent in quarantine per individual after 60 days. Cost and benefit are also evaluated at additional points in time (after 30, 90 or 120 days), see SM Section S2.5.4.

In all scenarios, we consider self-administered antigenic tests with turnaround time Δ_*w*_ = 15 minutes [20]. We assume the tests to have a 100% specificity, and a sensitivity *θ* which depends on the infectious compartment, with *θ*_*p*_ = 0.5, *θ*_*c*_ = 0.8, and *θ*_*sc*_ = 0.7 for the pre-symptomatic, clinical and sub-clinical compartments respectively. As sensitivity analysis, we consider in the SM the case of PCR tests with higher sensitivity and longer turnaround time (see SM Section S2.5.1).

### 2.3 Empirical contact data

We use high-resolution face-to-face empirical contacts data collected using wearable sensors in four different settings, two workplaces and two educational contexts: an office building, an hospital, a primary school and a high school. The data sets are publicly available on the website http://www.sociopatterns.org/datasets.

- The office data set (OD) gathers the contacts among 214 individuals, measured in an office building in France during two weeks in 2015 [40]. Individuals are divided in 12 departments with different sizes.
- The hospital data set (HD) describes the interaction among 42 health care workers (HCWs) and 29 patients in a hospital ward in Lyon, France, gathered during three days in 2010 [31]. HCWs are divided in three roles: nurses, doctors, and administrative staff.
- The primary school data set (PS) describes the contacts among 232 children and 10 teachers in a primary school in Lyon, France, during two days of school activity in 2009 [41]. The school is composed of 5 grades, each of them comprising 2 classes, for a total of 10 classes; there is a teacher for each class.
- The high school data set (CP) describes the contacts among 324 students of *classes préparatoires* in Marseille, France, during one week in 2013 [49]. These classes are located in high schools and are specific to the French schooling system: they gather students for 2-year studies at the end of the standard curriculum to prepare for entry exams at specific Universities. Students are grouped in 9 different classes, and classes are divided in three groups, each focusing on a specialization (mathematics and physics; physics, chemistry, engineering studies; biology).

Data sets are available as lists of contacts over time between anonymized individuals, with a classification by department (for OD), role (for HD) or class (for PS and CP), and in terms of students/teachers (for PS). From the raw data, we built the corresponding temporal contact networks, composed of nodes representing individuals, and links representing empirically measured proximity contacts occurring at a given time (see SM Section S1.1.1).

Figure 1b-e displays for each setting a graph of the links aggregated over one day for each data set (where the weight of a link between two individuals is given by the total contact time between them). The corresponding contact matrices representing the daily average density of interactions are shown in Figure 1f-i. In school settings and in offices, contacts occur preferentially within groups [41, 49, 40].

### 2.4 Data representations

The empirical data describes contacts at high resolution, giving temporally resolved information on who has been in contact with whom. These data can be aggregated into representations at different levels of detail, i.e., retaining only selected features of the empirical temporal contact network while aggregating over the others.

A first type of representations, which we denote by *individual-based representations*, preserve the empirical structure of the contact network (who has met whom).

- **Dynamical networks (DYN):** Contacts are aggregated into a different weighted graph for each successive time window of 15 minutes (The weight of a link between two nodes is given by the time in contact of the two corresponding individuals during this time window). This representation is the closest to the raw empirical data (that has a temporal resolution of 20 seconds), and will be considered as the reference against which the other representations will be compared.
- **Daily heterogeneous networks (dHET):** Contacts are aggregated into a different weighted graph for each of the *d*_*data*_ days of data collection. The weight *w*_*ij,d*_ of a link (*i, j*) on day *d* is given by the total contact time registered between *i* and *j* during the corresponding day.
- **Heterogeneous networks (HET):** Contacts measured during the whole data collection are aggregated into a single weighted network. The weight of a link (*i, j*) is given by the average daily contact time between *i* and *j*.

In a second type of representations, the *category-based representations*, we aggregate individuals into categories, corresponding to departments for the office data, to roles for the hospital data, and to classes in the school settings (and a category for teachers in the primary school data). Individuals belonging to a given category are considered as a priori equivalent. For each pair of categories *X* and *Y*, we summarize the interactions between individuals of these categories by the list of daily contact weights *D*_*XY*_ = {*w*_*ij,d*_|*i* ∈ *X, j* ∈ *Y, d* ∈ [1, *d*_*data*_]}. The average daily number of links between individuals of categories *X* and *Y* is *E*_*XY*_ = |*D*_*XY*_ |*/d*_*data*_, and the quantity *W*_*XY*_ = Σ_*i*∈*X,j*∈*Y,d*_ *w*_*ij,d*_*/d*_*data*_ gives the average daily total time in contact between individuals of categories *X* and *Y*. We define the three following data representations based on the concept of contact matrix [37]:

- **Contact matrix (CM):** Each individual from category *X* is connected with all individuals of category *Y* with a weight equal to *w*_*XY*_ = *W*_*XY*_ */*(*N*_*X*_*N*_*Y*_) (*N*_*X*_ is the number of individuals in category *X*; for *X* = *Y* we set *w*_*XX*_ = *W*_*XX*_*/*(*N*_*X*_ (*N*_*X*_ − 1)*/*2)). This representation only retains the average time spent in contact between members of given categories. For instance in the hospital data, *W*_*NUR,ADM*_ gives the total contact time between nurses and members of the administrative staff.
- **Contact matrix of bimodal distributions (CMB):** This representation, in addition, retains the information about the density of links between categories. To this aim, we create for each day a random graph assigning *E*_*XY*_ random links connecting individuals of categories *X* and *Y*. Each link is assigned a weight 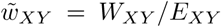 : contact durations between pairs of individuals depend only on their respective categories, but not all pairs of individuals are in contact. In the hospital data for instance, this representation retains the actual average daily number *E*_*NUR,ADM*_ of links between nurses and administrative staffs, and 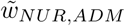 gives the average contact time on a link between a nurse and a member of the staff.
- **Contact matrix of distributions (CMD):** The CMD representation preserves the statistical heterogeneity of the daily contact durations between pairs of individuals. First, we create a random graph for each day as in the CMB case. The weight of each link between individuals of categories *X* and *Y* is then drawn from a negative binomial distribution, obtained by fitting the empirical distribution *D*_*XY*_ through a maximum likelihood procedure. In the same example as above, the representation not only retains *E*_*NUR,ADM*_, but it also uses the fitted distribution of all observed daily contact times between nurses and staff members.

Finally, we consider as a baseline a very coarse representation informed only by the total daily contact time:

- **Fully connected (FULL):** Individuals are all connected with each other. The weight of each link is equal to the daily contact time averaged over the whole data set *w* = Σ_*XY*_ *W*_*XY*_ */*(*N* (*N* − 1)*/*2), where *N* = Σ_*X*_ *N*_*X*_ is the total number of individuals.

Only the DYN representation retains information on the temporal evolution of contact activity along each day. However, we inform all other representations by the office or school hours and by the alternation of weekdays and week-ends, as reported in Table 3: no contacts occur outside of these hours nor on the week-ends.

**Table 3:**
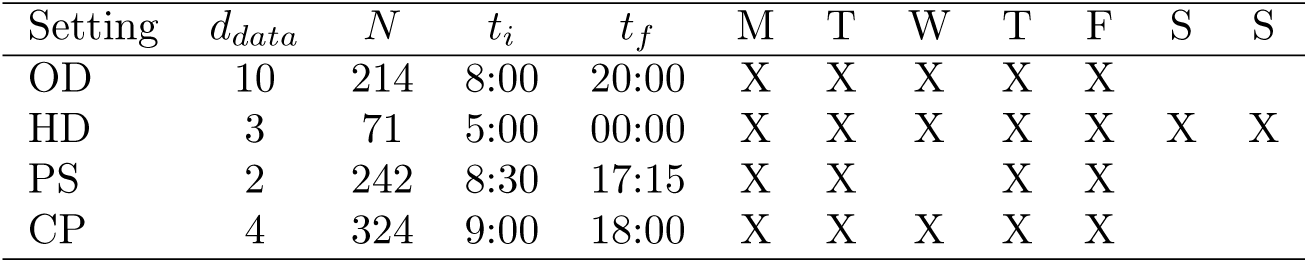
Number of days *d*_*data*_ of the data set, number of individuals *N*, initial hour (*t*_*i*_) and final hour (*t*_*f*_) of each day, and days of activity in each week (indicated with an X) for each setting.

| Setting | $d_{data}$ | $N$ | $t_i$ | $t_f$ | M | T | W | T | F | S | S |
| --- | --- | --- | --- | --- | --- | --- | --- | --- | --- | --- | --- |
| OD | 10 | 214 | 8:00 | 20:00 | X | X | X | X | X |  |  |
| HD | 3 | 71 | 5:00 | 00:00 | X | X | X | X | X | X | X |
| PS | 2 | 242 | 8:30 | 17:15 | X | X |  | X | X |  |  |
| CP | 4 | 324 | 9:00 | 18:00 | X | X | X | X | X |  |  |

### 2.5 Simulation setup

Simulations are initialized at a random time with one exposed individual chosen at random. Simulations then unfold in a stochastic way (see SM Section S1.2), with transmission events occurring, for each representation, along the contacts available in that representation of the data. To simulate the disease spreading on longer time scales than the available data (Table 3), copies of the initial data are repeated over time. Periodic introductions are considered to model infections from community. At regular intervals a susceptible individual in the considered setting is chosen at random and switched to the exposed compartment (see SM Section S1.2.5). To simulate a limited adherence to testing, the individuals accepting to perform tests are randomly chosen at the beginning of each simulation. Finally, we also explore the effect of initial immunity, simulated by the fact that a fraction of the population, randomly chosen at the start of each simulation, cannot be contaminated.

As discussed in [37, 38], simulations using a given rate of transmission *β* performed on different data representations yield different outcomes: less detailed representations tend to over-estimate the final size compared to the DYN representation [37], as they make more transmission paths available. Therefore, we fix a target basic reproductive number *R*_0_ in the absence of any control measures and starting with one random seed in an otherwise susceptible population, and calibrate for each representation the rate of transmission *β* needed to obtain the target *R*_0_ (see SM Section S1.3).

We consider two types of simulations. On the one hand, we study the dynamics of the spreading process in the absence of interventions (NT), starting from one random seed and with no introductions, and running simulations until no infectious individual is present in the population (Section 3.1). On the other hand, to evaluate NPIs, we consider in Section 3.2 simulations of a spread starting from one initial seed, with in addition bi-weekly introductions of exposed individuals. We simulate the spread for 60 days and compute the final epidemic size as well as the number of days that individuals spent in quarantine. Each result corresponds to a median over 2000 simulations, with bootstrapped confidence intervals (see SM Section S1.4).

## 3 Results

### 3.1 Unmitigated spread on different data representations

We present in the main text the results concerning the unmitigated spread in the office data set for *R*_0_ = 1.5 and *R*_0_ = 3, and we show in the SM Section S2.2 the results for the other data sets and both values of *R*_0_.

Figure 2 highlights differences and similarities between the processes taking place on different representations of the same data set. Figure 2a shows the distributions of the number of secondary cases resulting from one random seed, *R*_0,*i*_ (the basic reproductive number *R*_0_, which takes by construction the same value in all cases, being the average of this distribution), obtained on the various data representations. All distributions span a rather wide range of values, with events reaching almost four times the average. However, the curves exhibit distinct shapes depending on the type of representation. In the category-based representations, both small and large values of *R*_0,*i*_ are underestimated, i.e., both the probability that the spread never starts and the probability that super-spreading events occur. Fitting the distributions with negative binomials yields indeed values of the over-dispersion parameter *k* larger for the individual-based representations (≈0.5 for *R*_0_ = 3 in the office data set, see SM Section S2.2) than for the category-based ones (≈0.25 for CM, CMB, CMD and ≈0.22 for FULL, for *R*_0_ = 3 in the office data set, see SM Table S4). As shown in the SM Section S2.2, this effect is pronounced for OD and HD, but not in the school settings where individuals have a dense structure of contacts within each class.

**Figure 2:**
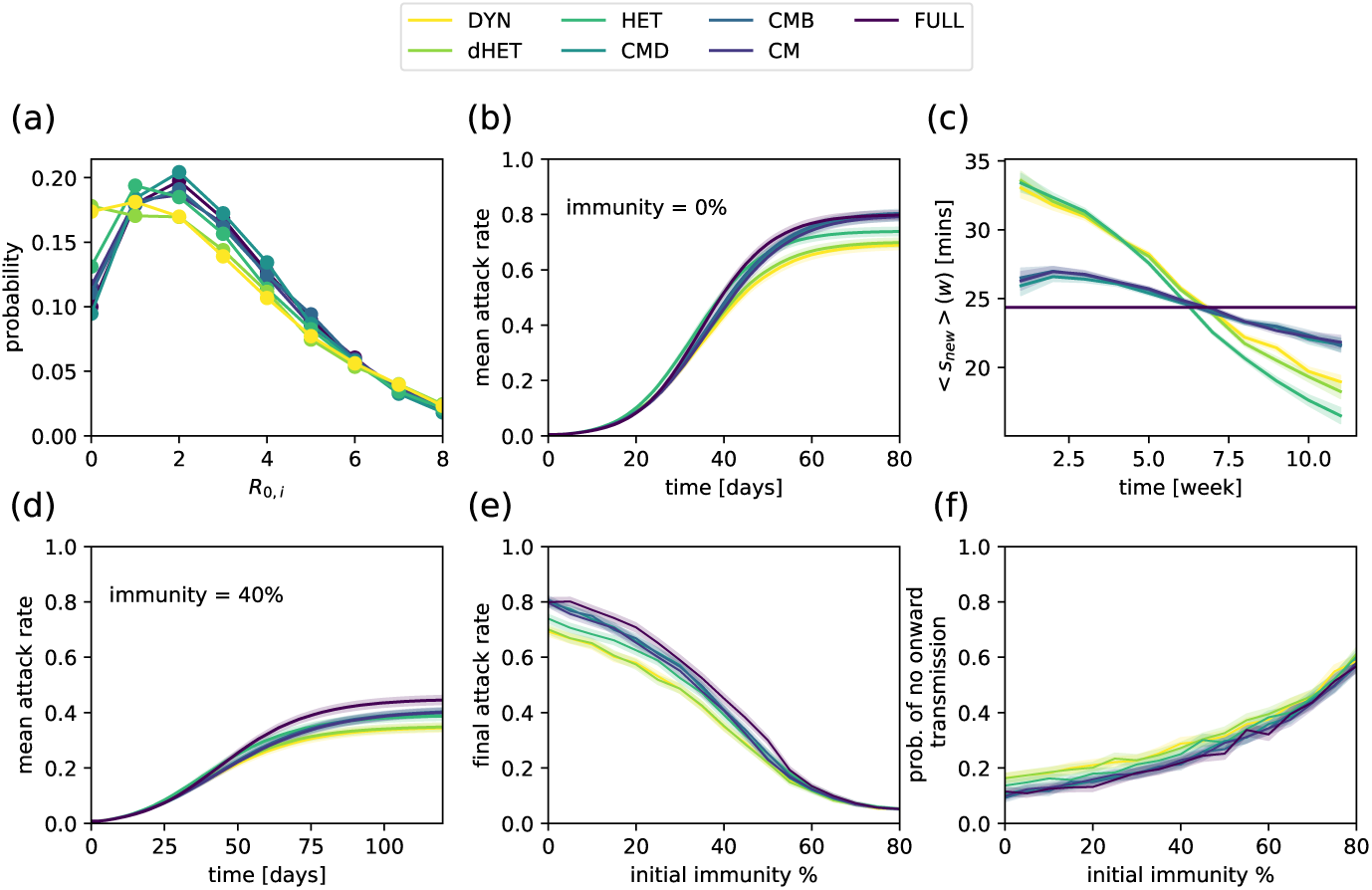
Spreading dynamics on different representations of the office data set, for *R*_0_ = 3.0, starting from a single initial exposed seed. (a) Distribution of the number of secondary infections produced by the initial seed. (b) Temporal evolution of the mean attack rate (fraction of individuals who have been infected), starting from one single exposed individual in an otherwise susceptible initial population. (c) Average strength (daily time in contact) of newly infected individuals infected in a given week vs. time. For individual-based representations, a cascade from more connected individuals to less connected ones is observed. The cascade is less pronounced for category-based representations. In panels (a,b,c), initial immunity is set to zero. (d) Temporal evolution of the mean attack rate, starting from one single exposed individual in a population with an initial immunity of 40%. (e) Mean final attack rate vs. initial immunity for different representations (here we compute the attack rate at the end of the simulation, i.e., when infectious individuals are no longer present). (f) Probability of no onward transmission (i.e., the initial seed does not infect any other individual) as a function of the initial immunity. Shaded areas correspond to the estimated error, obtained as a bootstrapped CI (see SM Section S1.4).

Another interesting difference between the two types of representations arises from the investigation of how the spread evolves within the population. Figure 2b shows the temporal evolution of the average fraction of individuals who have been infected at a given time for the various representations. The growth is slightly faster at short times for individual-based representations with respect to category-based ones, but saturates earlier and at smaller final epidemic sizes. These differences in dynamics can be understood by examining which nodes are infected at early and late stages of the spread. Indeed, as already discussed in [50], a spreading process on a network tends first to reach the most connected nodes, and a cascade towards the less connected nodes follows: the average number of neighbours of newly infected nodes decreases with time [50]. Here, as we deal with (weighted) contact networks in which heterogeneities concern contact times rather than numbers of neighbours [34], we show in Figure 2c the average daily strength *< s*_*new*_ *>* (*w*) of individuals who are infected and become exposed during week *w* (the strength *s* of an individual is the average daily time s/he spends in contact with other individuals). The cascading process from individuals with large *s* towards individuals with lower *s* is seen as a decreasing trend of *< s*_*new*_ *>* (*w*) for the individual-based representations. For the category-based representations, the cascade still exists, but the effect is weaker: all individuals within a category are equivalent, but some categories are more connected than others, so that some heterogeneity remains in the population. On the other hand, simulations using the FULL representation cannot show any such effect as all individuals are equivalent, and *< s*_*new*_ *>* (*w*) does not change with time. Overall, at early times the newly infected individuals are more connected in the individual-based representations than in category-based ones, leading to a faster spread. However, at later times, the tendency is inverted, with thus a slower spread on individual-based representations; moreover, as the remaining susceptible individuals tend to be less well connected, and as much less paths are available to reach them, the final epidemic size is also smaller. An additional difference is observed between the HET and the other individual-based representations: more causal propagation paths are present in the HET case (where the same network of contacts is present every day) than in the DYN and dHET cases, so that more nodes with smaller strength can be reached by the cascade and a larger epidemic size is obtained (as seen in Figure 2b).

Figure 2d-f explores the effect of an initial immunity of a part of the population. Figure 2d shows that differences in the dynamical evolution remain at intermediate immunity rates. Notably, the random immunization of nodes can change the ranking of final sizes obtained in different representations. Indeed, the category-based representations tend to make the networks of interactions more homogeneous than the individual-based ones, and random immunization is less efficient in heterogeneous than in homogeneous networks [51], so that the final epidemic size can become smaller for the category-based representations (Figure 2d). When increasing the initial immunity, the effect on the final attack rate is similar for all representations. In particular, the mean attack rate decreases to small values at approximately the same fraction of immune individuals (Figure 2e), and the probability of rapid extinction of the spread becomes similar for all representations (Figure 2f).

### 3.2 Robustness of the evaluation of NPIs

We show here the results of simulations implementing NPIs for *R*_0_ = 1.5, and present additional results and sensitivity analysis in the SM Sections S2.3-S2.5.

Figure 3 first shows the impact of the baseline intervention, ST (testing and isolation of symptomatic cases), in different settings and when using different data representations when *R*_0_ = 1.5. Panel (a) displays the effective reproductive number when ST is implemented (short term dynamics), compared with the baseline no test scenario (NT), and panel (b) the median attack rate after 60 days, comparing NT and ST. Several observations are in order. Even at fixed *R*_0_ and with no interventions, the median attack rate after 60 days depends on the context. For all representations, it is much higher for the HD data set (Figure 3b). Most representations also yield the same ranking of contexts as a function of the attack rate, with the exception of the FULL representation, which strongly overestimates the attack rate in the school contexts (where the structure of the population in groups is also the strongest). The impact of the ST protocol also differs depending on context, with a smaller effect in the school settings, for all representations, both in terms of decreasing the effective reproductive number (Figure 3a) and the final attack rate (Figure 3b). This could be explained by the strong density of links within classes and the fact that the probability of developing clinical symptoms is smaller for children and adolescents (in addition, the probability of symptomatic detection is smaller for children).

**Figure 3:**
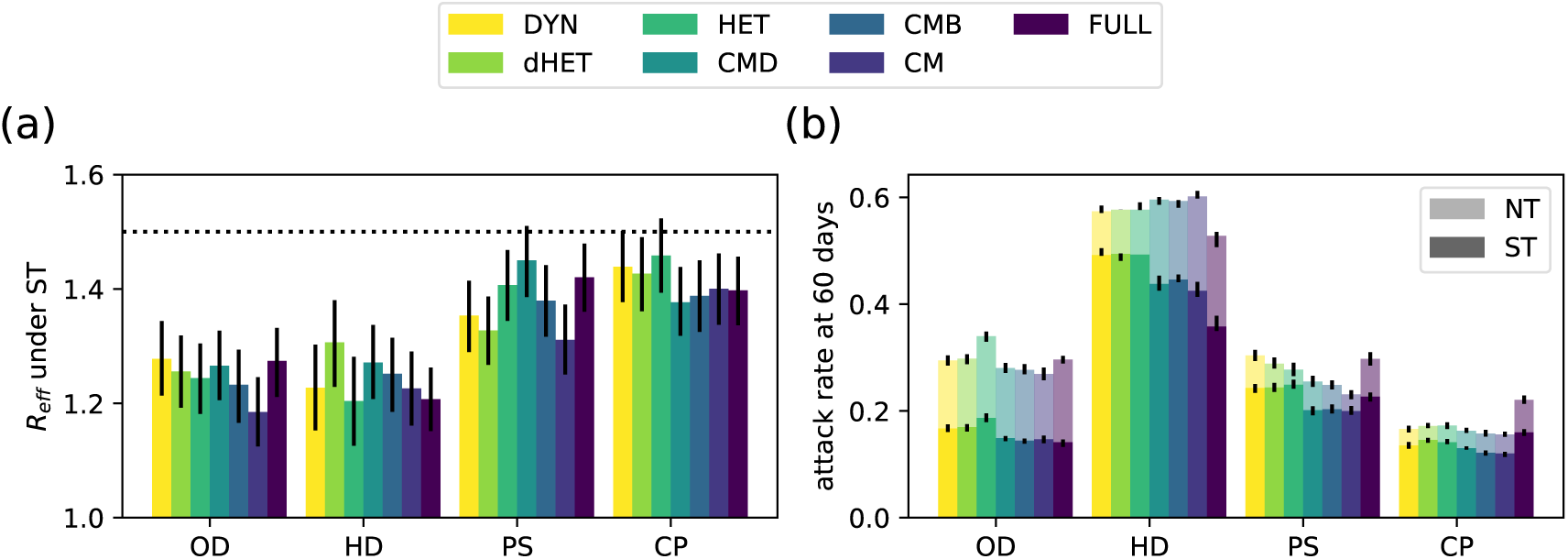
Effect of symptomatic testing (ST) in all settings and for all representations, for *R*_0_ = 1.5. (a) Effective reproductive number *R*_*eff*_ with ST implemented, measured as the average number of secondary infections caused by an initial infected seed. The horizontal dashed line corresponds to *R*_*eff*_ = 1.5 (corresponding to no interventions). (b) Median attack rate in each setting, without interventions (NT) and with ST in place.

As testing and isolation of symptomatic individuals is the minimal strategy that can be put in place, we now focus on a comparison of all protocols with respect to ST. We present the results for the OD and PS data sets in Figure 4, and show the results for other data sets in the SM Section S2.3, as well as additional values of the protocols’ parameters. Figure 4a-b shows the reduction in the median epidemic size after 60 days for several protocols, with respect to the ST one, with protocols ranked in order of increasing such reduction when simulated on the DYN data representation. Strikingly, even if the precise values of the efficacy of each protocol depend slightly on the data representation used in the simulations, the ranking of protocols remains almost always the same, both for benefits (Figure 4a,b) and costs (Figure 4c,d). In particular, telework in the offices is particularly efficient, as it reduces the number of contacts of all individuals [19], and reactive strategies at school are less efficient than regular testing [20], due to the fact that reactive strategies are implemented only upon detection of symptomatic cases: silent transmission by pre-symptomatic and asymptomatic infectious to other classes is very likely to occur before the strategies can contain the spread, as shown in [20]. These conclusions can be reached whatever the data representation used to perform the simulations. Note that the robustness of the ranking with respect to the representation is very strong but not perfect: if two protocols yield average efficacy values that are very close, one can seem slightly better for one representation and slightly worse for another; moreover, some exceptions can be observed, such as the case of the FULL representation, which strongly underestimates the efficacy of the reactive testing (rT) protocol. Figure 4e-f show that the impact of a protocol on the distributions of epidemic sizes is also similar across representations: here, regular testing yields a strong reduction of the probability of having a large epidemic size and a much higher peak at small sizes. We also show in the SM Section S2.3 how, when two protocols have similar efficacies, the resulting distributions of epidemic sizes are also very similar, and that this similarity holds across representations.

**Figure 4:**
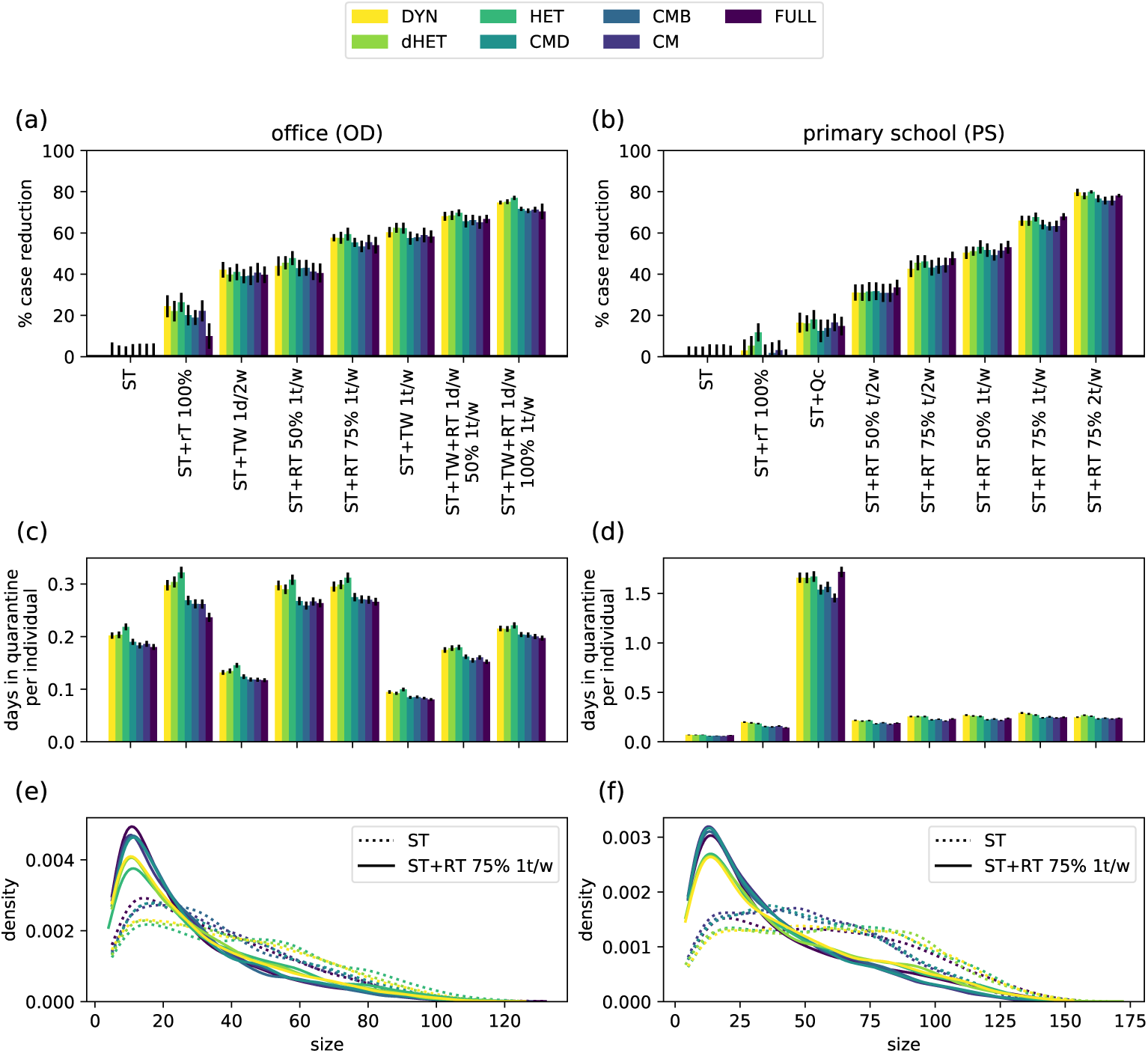
Evaluation of several NPIs in offices (OD) and primary school (PS) settings, for *R*_0_ = 1.5 and simulations performed using different data representations. (a,b) Efficacy of NPIs in offices and primary school, sorted by increasing order of efficacy in the DYN representation. Efficacy is defined as the relative reduction in median size compared with symptomatic (ST) testing alone, after a period of 60 days. (c,d) Average number of days in quarantine per individual under different protocols. (e-f) Epidemic size distributions for the baseline protocol ST (dotted lines), and for weekly regular testing with 75% adherence (continuous line).

We illustrate these results further in Figure 5, where we investigate the question of the adherence to regular testing needed in offices to obtain the same efficacy as telework, for a given testing frequency (Figure 5a). Although the value of the median size reduction obtained by TW slightly depends on the data representation (TW 1t/w yields a 59±3% and 60±3% reduction for CM and DYN representations, respectively), we can estimate that RT with the same frequency becomes as efficient as TW for adherence values that remain similar across data representations, ranging from 84% (CM representation) to 81% (DYN representation). Figure 5b considers instead the comparison between the regular testing and the class quarantine protocol ST+Q_*c*_: the estimation of the adherence needed for ST+RT*α* to become more efficient than ST+Q_*c*_ is also consistent across data representations. Another interesting point concerns the effect of increasing the number of tests, either by increasing adherence or by increasing frequency, within the RT protocol. First, the increase in efficacy faces diminishing returns: Figure 5c-d illustrates this by showing the average size reduction per test for the RT protocol with frequency once per week and adherence 25%, and comparing it with the additional size reduction per test obtained for twice the number of tests, obtained either by doubling the adherence at the same frequency, or by doubling the frequency at the same adherence. Second, and as already noted in [20] with simulations on the DYN representation of a school data set, increasing adherence has a bigger impact than an increase in frequency (at equal additional number of tests). We show in the SM Section S2.3.3 that this property holds in all settings, and for all data representations.

**Figure 5:**
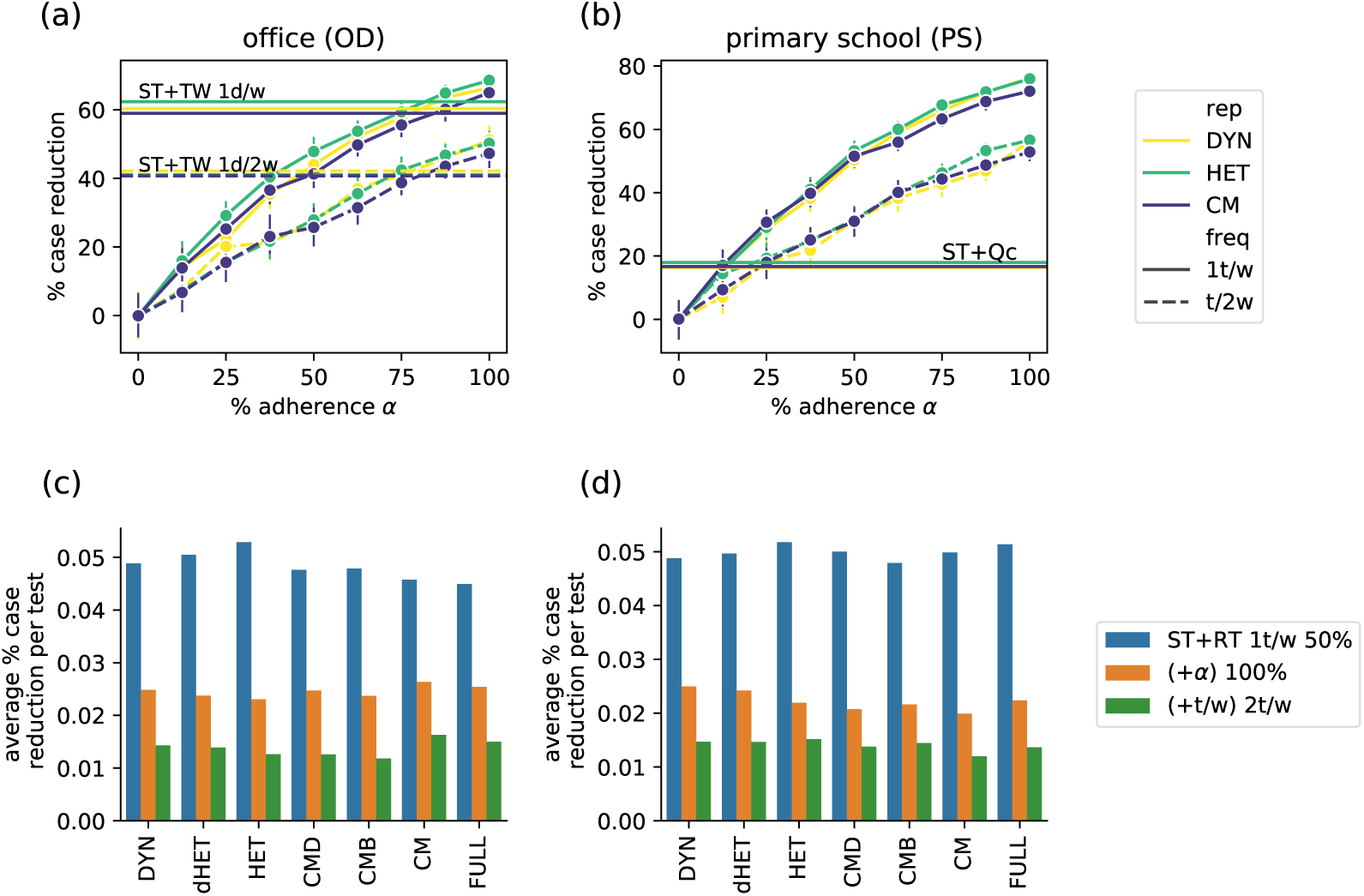
Effect of increasing adherence and frequency in regular testing protocols. (a) Effect of the adherence *α* for a given frequency (once per week or every two weeks) in the ST+RT*α* protocol for the office data set and *R*_0_ = 1.5, compared with telework (TW), for several data representations. Horizontal lines correspond to the performance of TW at the same frequencies. (b) Effect of the adherence *α* for a given frequency (once per week or every two weeks) in the ST+RT*α* protocol, compared with the class quarantine protocol ST+Q_*c*_, for the school data set and *R*_0_ = 1.5. Horizontal lines correspond to the ST+Q_*c*_ protocol. (c,d) Effect of improving adherence or frequency, for *R*_0_ = 1.5 for OD (c) and PS (d). We consider ST+RT at frequency once per week and *α* = 50%, and we measure the average size reduction (w.r.t. ST) per test (in blue), and the additional size reduction per additional test when doubling the adherence (in orange), and when doubling the frequency (in green).

We finally examine in the SM Section S2.3.2 the impact of the reproductive number *R*_0_. As also observed in [20, 47], the efficacy of each protocol depends in a non-monotonic way on *R*_0_. At small *R*_0_, even the basic ST protocol leads to small epidemic sizes, so that additional protocols have a limited impact. At very large *R*_0_ instead, even the best protocols reach their limits and the spread cannot be well mitigated. These arguments hold for any data representation, and we indeed observe this non-monotonicity for all data representations. However, the optimal range of *R*_0_ depends on the data representation, with an overestimation of the optimal *R*_0_ for the category-based representations. Moreover, the differences between the efficacy values of a given protocol by using different data representations become larger at large *R*_0_, with an overestimation of the efficacy by category-based representations.

Different protocols have different efficacies but also different costs, which need to be taken into account in decision making processes. We thus compare in Figure 4c-d the cost of each protocol simulated on each data representation, computed as the average number of days spent in quarantine per node. As for the efficacy, the precise evaluation of the cost depends on the data representation, but the ranking of protocols according to their cost does not. In particular, regular testing at school avoids a large fraction of the number of days of class lost, with respect to reactive class closures. In the offices, regular testing is more costly than telework, as the latter simply decreases the number of contacts without quarantining individuals.

Overall, Figure 4 indicates that a coherent picture of the relative efficacy and cost of different protocols is obtained when using different representations of the data in the numerical simulations, even if quantitative differences in the precise evaluation are observed. Additional results shown in the SM Section S2.5 indicate that these conclusions are robust with respect to changes in disease and protocol parameters. We also explore in the SM Section S2.4 the combined effect of NPIs and vaccination: vaccination reduces the final epidemic size even in the absence of NPIs or for the basic ST protocol, and decreases the costs in terms of quarantines. This effect can be assessed using any data representation. Moreover, even in the presence of vaccination, all data representations agree on the ranking of protocols according to their efficacy or cost, and on the fact that NPIs remain very valuable additional tools at intermediate vaccination coverages [20, 43].

## 4 Discussion

We used high-resolution contact data sets to build aggregated representations and evaluate how loss of resolution informing epidemic models can influence the evaluation of prevention and control strategies. Numerical simulations of a model for the spread of SARS-CoV-2 in educational and professional contexts show that detailed representations are needed to correctly account for over-dispersion of reproduction numbers and for a precise evaluation of the efficacy and costs of each strategy. However, coarse representations containing only very summarized information are good enough to rank protocols, and thus to provide insights on better options given the context.

Models offer a unique opportunity to evaluate strategies for prevention and control of epidemics, anticipating their expected advantage and costs associated to inform public health decisions. Depending on the context and the question to be addressed, models need to integrate an accurate description of the population under study and of the contacts along which disease transmission occurs. In recent years, the increasing availability of data sets describing contacts between individuals has made it possible to devise models exposing the complexity of human interactions in terms of number of contacts, repeated contacts, frequency, duration, etc. For instance, models integrating data describing interactions with high temporal and spatial resolutions can be used to design and study measures tailored to specific contexts such as schools, where repetition of contacts because of friendships and structural organization of contacts due to classes impact the resulting epidemic dynamics [14, 46, 21, 20]. Complex models are however data hungry, and detailed data are not always available. Moreover, data sets in specific settings may provide a narrow vision of the interaction patterns occurring in those contexts and may be difficult to generalize. By loosing some of these specificities, aggregated representations may become more generally applicable.

Our results show that some differences emerge in the disease spread simulated on different data representations, even when calibrating the simulations to yield the same basic reproductive numbers. In particular, category-based representations tend to underestimate the over-dispersion of the distribution of the reproductive number, and could thus lead to difficulties in correctly estimating the role of super-spreading events. This is in line with recent results highlighting the role of contact heterogeneities in super-spreading [36]. As they ignore individual differences, these representations cannot inform strategies targeted towards specific individuals, they are also less able to describe the cascading of a spread from individuals with a high connectivity to less well connected ones [50], and lead to an overestimation of the final epidemic size [37].

The picture is more complex when dealing with the evaluation of control protocols. On the one hand, the ranking of protocols according to their efficacy or their cost does not depend on the data representation. The picture of which protocol is most efficient in each context remains coherent. When a protocol depends on several parameters, the information on which parameter is the most important to act upon is also coherent across data representations (e.g., increasing adherence for regular testing protocols has a larger impact than increasing frequency, at given number of tests). It is even possible to use coarse data representations to quantify the adherence needed for the regular testing to become more efficient than e.g. telework or class quarantine. On the other hand, using various data representations can lead to quantitative differences in the precise values of benefit and cost. This can be a limitation for coarse representations when, despite a coherent ranking in efficiency of protocols, decisions require accuracy in the estimate of the benefit/cost – for example, to define a minimum benefit that would trigger the application of the measure. Such decisions should thus take into account an inherent uncertainty in the model outcomes due to the limited information contained in the data.

In the contexts we considered, we found that regular testing with high enough adherence is a very efficient strategy allowing to limit spread in school contexts while minimizing the number of lost school-days, confirming prior works [20, 52, 21]. In offices, telework is also very efficient [19]. Reactive class closure or reactive testing instead have limited efficacy, and class closure naturally leads to a strong cost in terms of lost school-days. The robustness of such results across data representations is explained by the fact that these NPIs reduce the epidemic size through mechanisms that do not depend on the data description. Indeed, symptomatic testing, class quarantines and reactive testing are reactive measures triggered by the detection of cases. However, the infectiousness of pre-symptomatic and asymptomatic individuals strongly limits the efficacy of such measures: for instance, due to the resulting silent propagation, many other classes can already have been reached by the infection when one class is closed upon the detection of a case at school [20]. In contrast, regular testing is a proactive approach that allows to detect also pre-symptomatic and asymptomatic cases. Telework on the other side simply reduces the time in contact, thus effectively reducing the probability of contagion events, whatever the data representation. Overall, our results support the use of even coarse representations of the interactions between individuals in settings such as schools or workplaces when evaluating NPIs and potentially choosing between possible protocols.

Individual data such as the ones used in this study across different settings are rarely available. Moreover, the existing data sets are each specific to a context and potentially to the time of the data collection campaign. In emergency situations or during a crisis such as the current pandemic, gathering such data in real time encounters many challenges, and more coarse-grained representations are generally opted for. Indeed, summarized data is more accessible, and can be enriched by some robust statistical features of contact data, such as the heterogeneities in contact durations [29, 37, 34], as in the CMD data representation [37]: such information could be reconstructed from limited data describing a given context [37, 42]. In particular, division of a population into categories with e.g. different mixing patterns and/or schedules can be performed from limited information such as the existence of classes in a school or of departments in offices. A population can also be separated in groups according to an expected diversity of behaviours, as for instance in [43] that singles out the group of “more social” students in a US campus as the ones belonging to fraternities and show that targeted testing of this category can be an efficient strategy.

Our work comes with limitations that are worth discussing. First, the data we used describe contacts collected during only few days. Here, we have used the simplest method of repeating the data set in order to simulate the contacts in the population during an extended time, while contacts are not repeated identically in the real world. However, the simulations performed in [20] used different ways of artificially extending the data duration and found no differences in the results. The settings we have considered are also relatively small, but represent the state of the art in terms of data describing interactions between individuals, and have very different structural and temporal properties. These settings are moreover very important ones in practice, for which protocols need to be tailored and evaluated. Further work could also consider data at larger scale or with more heterogeneous contact structures (e.g., with categories of very different sizes, or with more heterogeneous distributions of the numbers of neighbours of each node) if they become available, or synthetic data representing larger contexts. Second, we used a rather simple coupling with the community, through regular introduction of cases, as the data we considered do not include contacts happening outside of the studied context. Even though our results are robust with respect to variations in the frequency of introductions, it would be desirable to inform the model with empirical data on the contacts that individuals have with members of the community, or with one another outside of school. Third, we have here considered one specific infectious disease. However, our results are robust with respect to variations in the basic reproductive number, initial immunity, and the impact of vaccination. Moreover, this disease is of particular interest both practically and theoretically, as the pre-symptomatic and asymptomatic transmissions make it necessary to go beyond the usual reactive strategies and to evaluate a range of protocols.

Finally, our modelling approaches deal with the interactions between individuals but do not address the issue of individual heterogeneities with respect to the disease transmission (beyond the differences between children, adolescents, adults), such as heterogeneous infectious periods [53] or heterogeneous rates of transmission [54], nor with respect to potential changes of behaviour depending on the epidemic situation [55]. An interesting extension of this work would be to consider situations where these differences between individuals are correlated with their contact behaviour: to take into account such correlations, one would need to go beyond the category-based representations we have considered here, allowing heterogeneous properties within each category, in the spirit of degree-corrected stochastic block models [56].

## Data Availability

All data produced in the present study are available upon reasonable request to the authors

## Acknowledgments

This study was partially funded by: ANR projects COSCREEN (ANR-21-CO16-0005) and DATARE-DUX (ANR-19-CE46-0008-03); ANRS-MIE project EMERGEN (ANRS0151); EU H2020 grants MOOD (H2020-874850) and RECOVER (H2020-101003589); EU HORIZON grant VERDI (101045989); REACTing COVID-19 grant.

## References

[1] Roy M Anderson and Robert M May. Infectious diseases of humans: dynamics and control. Oxford university press, 1992.

[2] Matt J Keeling and Pejman Rohani. Modeling infectious diseases in humans and animals. Princeton University Press, 2011.

[3] D. Balcan, V. Colizza, B. Gonçalves, H. Hu, J.J. Ramasco, and A. Vespignani. Multiscale mobility networks and the spatial spreading of infectious diseases. Proc Natl Acad Sci USA, 106:21484–21489, 2009.

[4] V. Colizza, A. Barrat, M. Barthelemy, A.-J. Valleron, and A. Vespignani. Modeling the worldwide spread of pandemic influenza: Baseline case and containment interventions. PLOS Medicine, 4(1):e13, 2007.

[5] Neil M. Ferguson, Derek A. T. Cummings, Christophe Fraser, James C. Cajka, Philip C. Cooley, and Donald S. Burke. Strategies for mitigating an influenza pandemic. Nature, 442(7101):448–452, 2006.

[6] Juliette Stehlé, Nicolas Voirin, Alain Barrat, Ciro Cattuto, Vittoria Colizza, Lorenzo Isella, Corinne Régis, Jean-Francois Pinton, Nagham Khanafer, Wouter Van den Broeck, and Philippe Vanhems. Simulation of an seir infectious disease model on the dynamic contact network of conference attendees. BMC Medicine, 9(1):87, 2011.

[7] M. Tizzoni, P. Bajardi, C. Poletto, J.J. Ramasco, D. Balcan, B. Goncalves, N. Perra, V. Colizza, and A. Vespignani. Real-time numerical forecast of global epidemic spreading: case study of 2009 a/h1n1pdm. BMC Medicine, 10:165, 2012.

[8] Matteo Chinazzi, Jessica T. Davis, Marco Ajelli, Corrado Gioannini, Maria Litvinova, Stefano Merler, Ana Pastore y Piontti, Kunpeng Mu, Luca Rossi, Kaiyuan Sun, Cécile Viboud, Xinyue Xiong, Hongjie Yu, M. Elizabeth Halloran, Ira M. Longini, and Alessandro Vespignani. The effect of travel restrictions on the spread of the 2019 novel coronavirus (covid-19) outbreak. Science, 368(6489):395–400, 2020.

[9] Giulia Pullano, Francesco Pinotti, Eugenio Valdano, Pierre-Yves Boëlle, Chiara Poletto, and Vittoria Colizza. Novel coronavirus (2019-ncov) early-stage importation risk to europe, january 2020. Eurosurveillance, 25(4), 2020.

[10] Laura Di Domenico, Giulia Pullano, Chiara E Sabbatini, Pierre-Yves Boëlle, and Vittoria Colizza. Impact of lockdown on covid-19 epidemic in île-de-france and possible exit strategies. BMC medicine, 18(1):1–13, 2020.

[11] Moritz U. G. Kraemer, Chia-Hung Yang, Bernardo Gutierrez, Chieh-Hsi Wu, Brennan Klein, David M. Pigott, null null, Louis du Plessis, Nuno R. Faria, Ruoran Li, William P. Hanage, John S. Brownstein, Maylis Layan, Alessandro Vespignani, Huaiyu Tian, Christopher Dye, Oliver G. Pybus, and Samuel V. Scarpino. The effect of human mobility and control measures on the covid-19 epidemic in China. Science, 368(6490):493–497, 2020.

[12] Juanjuan Zhang, Maria Litvinova, Yuxia Liang, Yan Wang, Wei Wang, Shanlu Zhao, Qianhui Wu, Stefano Merler, Cécile Viboud, Alessandro Vespignani, et al. Changes in contact patterns shape the dynamics of the COVID-19 outbreak in China. Science, 368(6498):1481–1486, 2020.

[13] Laura Di Domenico, Chiara E Sabbatini, Giulia Pullano, Daniel Lévy-Bruhl, and Vittoria Colizza. Impact of january 2021 curfew measures on sars-cov-2 b.1.1.7 circulation in france. Eurosurveillance, 26(15), 2021.

[14] Valerio Gemmetto, Alain Barrat, and Ciro Cattuto. Mitigation of infectious disease at school: Targeted class closure vs school closure. BMC Infectious Diseases, 14(1):1–10, 2014.

[15] Constanze Ciavarella, Laura Fumanelli, Stefano Merler, Ciro Cattuto, and Marco Ajelli. School closure policies at municipality level for mitigating influenza spread: a model-based evaluation. BMC Infectious Diseases, 16(1):576, 2016.

[16] Joel R Koo, Alex R Cook, Minah Park, Yinxiaohe Sun, Haoyang Sun, Jue Tao Lim, Clarence Tam, and Borame L Dickens. Interventions to mitigate early spread of sars-cov-2 in singapore: a modelling study. The Lancet Infectious Diseases, 20(6):678–688, 2020.

[17] Adam J Kucharski, Petra Klepac, Andrew JK Conlan, Stephen M Kissler, Maria L Tang, Hannah Fry, Julia R Gog, W John Edmunds, Jon C Emery, Graham Medley, et al. Effectiveness of isolation, testing, contact tracing, and physical distancing on reducing transmission of sars-cov-2 in different settings: a mathematical modelling study. The Lancet Infectious Diseases, 20(10):1151–1160, 2020.

[18] David R. M. Smith, Audrey Duval, Koen B. Pouwels, Didier Guillemot, Jérôme Fernandes, Bich-Tram Huynh, Laura Temime, Lulla Opatowski, and on behalf of the AP-HP/Universities/Inserm COVID-19 research collaboration. Optimizing covid-19 surveillance in long-term care facilities: a modelling study. BMC Medicine, 18(1):386, 2020.

[19] Simon Mauras, Vincent Cohen-Addad, Guillaume Duboc, Max Dupré la Tour, Paolo Frasca, Claire Mathieu, Lulla Opatowski, and Laurent Viennot. Mitigating covid-19 outbreaks in workplaces and schools by hybrid telecommuting. PLOS Computational Biology, 17(8):1–24, 08 2021.

[20] Elisabetta Colosi, Giulia Bassignana, Diego Andrés Contreras, Canelle Poirier, Simon Cauchemez, Yazdan Yazdanpanah, Bruno Lina, Arnaud Fontanet, Alain Barrat, and Vittoria Colizza. Selftesting and vaccination against covid-19 to minimize school closure. Lancet Inf. Diseases, in press, 2022.

[21] Ryan Seamus McGee, Julian R. Homburger, Hannah E. Williams, Carl T. Bergstrom, and Alicia Y. Zhou. Model-driven mitigation measures for reopening schools during the covid-19 pandemic. Proceedings of the National Academy of Sciences, 118(39), 2021.

[22] Quan-Hui Liu, Juanjuan Zhang, Cheng Peng, Maria Litvinova, Shudong Huang, Piero Poletti, Filippo Trentini, Giorgio Guzzetta, Valentina Marziano, Tao Zhou, Cecile Viboud, Ana I. Bento, Jiancheng Lv, Alessandro Vespignani, Stefano Merler, Hongjie Yu, and Marco Ajelli. Model-based evaluation of alternative reactive class closure strategies against covid-19. Nature Communications, 13(1):322, 2022.

[23] J. Mossong, N. Hens, M. Jit, P. Beutels, K. Auranen, R. Mikolajczyk, M. Massari, S. Salmaso, G. Scalia Tomba, J. Wallinga, J. Heijne, M. Sadkowska-Todys, M. Rosinska, and W.J. Edmunds. Social Contacts and Mixing Patterns Relevant to the Spread of Infectious Diseases. PLoS Medicine, 5(3):e74, 2008.

[24] Romualdo Pastor-Satorras, Claudio Castellano, Piet Van Mieghem, and Alessandro Vespignani. Epidemic processes in complex networks. Rev. Mod. Phys., 87:925–979, Aug 2015.

[25] K. Eames, S. Bansal, S. Frost, and S. Riley. Six challenges in measuring contact networks for use in modelling. Epidemics, 10:72–77, 2015. Challenges in Modelling Infectious DIsease Dynamics.

[26] Naoki Masuda and Petter Holme, editors. Temporal Network Epidemiology. Springer, Singapore, 2017.

[27] J. Wallinga, P. Teunis, and M. Kretzschmar. Using data on social contacts to estimate age-specific transmission parameters for respiratory-spread infectious agents. Am J Epidemiol, 164:936–944, 2006.

[28] Leon Danon, Jonathan M. Read, Thomas A. House, Matthew C. Vernon, and Matt J. Keeling. Social encounter networks: characterizing great britain. Proceedings of the Royal Society B: Biological Sciences, 280(1765), 2013.

[29] Ciro Cattuto, Wouter Van den Broeck, Alain Barrat, Vittoria Colizza, Jean-François Pinton, and Alessandro Vespignani. Dynamics of person-to-person interactions from distributed rfid sensor networks. PLoS ONE, 5(7):e11596, 07 2010.

[30] Marcel Salathé, Maria Kazandjieva, Jung Woo Lee, Philip Levis, Marcus W. Feldman, and James H. Jones. A high-resolution human contact network for infectious disease transmission. Proceedings of the National Academy of Sciences, 107(51):22020–22025, 2010.

[31] Philippe Vanhems, Alain Barrat, Ciro Cattuto, Jean François Pinton, Nagham Khanafer, Corinne Régis, Byeul a. Kim, Brigitte Comte, and Nicolas Voirin. Estimating potential infection transmission routes in hospital wards using wearable proximity sensors. PloS one, 8(9), 2013.

[32] Arkadiusz Stopczynski, Vedran Sekara, Piotr Sapiezynski, Andrea Cuttone, Mette My Madsen, Jakob Eg Larsen, and Sune Lehmann. Measuring large-scale social networks with high resolution. PLoS ONE, 9(4):e95978, 04 2014.

[33] Damon J. A. Toth, Molly Leecaster, Warren B. P. Pettey, Adi V. Gundlapalli, Hongjiang Gao, Jeanette J. Rainey, Amra Uzicanin, and Matthew H. Samore. The role of heterogeneity in contact timing and duration in network models of influenza spread in schools. Journal of The Royal Society Interface, 12(108), 2015.

[34] Alain Barrat and Ciro Cattuto. Face-to-Face Interactions, pages 37–57. Springer International Publishing, Cham, 2015.

[35] Sally Blower and Myong-Hyun Go. The importance of including dynamic social networks when modeling epidemics of airborne infections: does increasing complexity increase accuracy? BMC medicine, 9(1):1–3, 2011.

[36] Zachary Susswein and Shweta Bansal. Characterizing superspreading of sars-cov-2 : from mechanism to measurement. medRxiv, 2020.

[37] Anna Machens, Francesco Gesualdo, Caterina Rizzo, Alberto E. Tozzi, Alain Barrat, and Ciro Cattuto. An infectious disease model on empirical networks of human contact: bridging the gap between dynamic network data and contact matrices. BMC Infectious Diseases, 13(1):1–18, 2013.

[38] Alberto Aleta, Guilherme Ferraz de Arruda, and Yamir Moreno. Data-driven contact structures: From homogeneous mixing to multilayer networks. PLoS computational biology, 16(7):e1008035, 2020.

[39] Livio Bioglio, Mathieu Génois, Christian L. Vestergaard, Chiara Poletto, Alain Barrat, and Vittoria Colizza. Recalibrating disease parameters for increasing realism in modeling epidemics in closed settings. BMC Infectious Diseases, 16(1):1–15, 2016.

[40] Mathieu Génois and Alain Barrat. Can co-location be used as a proxy for face-to-face contacts? EPJ Data Science, 7(1):1–18, 2018.

[41] Juliette Stehlé, Nicolas Voirin, Alain Barrat, Ciro Cattuto, Lorenzo Isella, Jean-François Pinton, Marco Quaggiotto, Wouter Van den Broeck, Corinne Régis, Bruno Lina, et al. High-resolution measurements of face-to-face contact patterns in a primary school. PloS one, 6(8):e23176, 2011.

[42] Timo Smieszek and Marcel Salathé. A low-cost method to assess the epidemiological importance of individuals in controlling infectious disease outbreaks. BMC medicine, 11(1):1–8, 2013.

[43] Peter I. Frazier, J. Massey Cashore, Ning Duan, Shane G. Henderson, Alyf Janmohamed, Brian Liu, David B. Shmoys, Jiayue Wan, and Yujia Zhang. Modeling for COVID-19 college reopening decisions: Cornell, a case study. Proceedings of the National Academy of Sciences, 119(2), 2022.

[44] Christopher M. Baker, Iadine Chades, Jodie McVernon, Andrew P. Robinson, and Howard Bondell. Optimal allocation of pcr tests to minimise disease transmission through contact tracing and quarantine. Epidemics, 37:100503, 2021.

[45] Laura Di Domenico, Giulia Pullano, Chiara E Sabbatini, Pierre-Yves Boëlle, and Vittoria Colizza. Modelling safe protocols for reopening schools during the covid-19 pandemic in france. Nature communications, 12(1):1–10, 2021.

[46] Jana Lasser, Johannes Sorger, Lukas Richter, Stefan Thurner, Daniela Schmid, and Peter Klimek. Assessing the impact of sars-cov-2 prevention measures in austrian schools using agent-based simulations and cluster tracing data. Nature Communications, 13(1):554, 2022.

[47] A. Barrat, C. Cattuto, M. Kivelä, S. Lehmann, and J. Saramäki. Effect of manual and digital contact tracing on COVID-19 outbreaks: A study on empirical contact data. Journal of the Royal Society Interface, 18(178), 2021.

[48] Jesús A. Moreno López, Beatriz Arregui García, Piotr Bentkowski, Livio Bioglio, Francesco Pinotti, Pierre-Yves Boëlle, Alain Barrat, Vittoria Colizza, and Chiara Poletto. Anatomy of digital contact tracing: Role of age, transmission setting, adoption, and case detection. Science Advances, 7(15):eabd8750, 2021.

[49] Rossana Mastrandrea, Julie Fournet, and Alain Barrat. Contact patterns in a high school: a comparison between data collected using wearable sensors, contact diaries and friendship surveys. PloS one, 10(9):e0136497, 2015.

[50] Marc Barthélemy, Alain Barrat, Romualdo Pastor-Satorras, and Alessandro Vespignani. Dynamical patterns of epidemic outbreaks in complex heterogeneous networks. Journal of theoretical biology, 235(2):275–288, 2005.

[51] A. Barrat, M. Barthélemy, and A. Vespignani. Dynamical processes on complex networks. Cambridge University Press, Cambridge, 2008.

[52] Ryan Seamus McGee, Julian R Homburger, Hannah E Williams, Carl T Bergstrom, and Alicia Y Zhou. Proactive covid-19 testing in a partially vaccinated population. medRxiv, 2021.

[53] Alexandre Darbon, Davide Colombi, Eugenio Valdano, Lara Savini, Armando Giovannini, and Vittoria Colizza. Disease persistence on temporal contact networks accounting for heterogeneous infectious periods. Royal Society open science, 6(1):181404, 2019.

[54] Hui Yang, Ming Tang, and Thilo Gross. Large epidemic thresholds emerge in heterogeneous networks of heterogeneous nodes. Scientific reports, 5(1):1–12, 2015.

[55] Sebastian Funk, Shweta Bansal, Chris T. Bauch, Ken T.D. Eames, W. John Edmunds, Alison P. Galvani, and Petra Klepac. Nine challenges in incorporating the dynamics of behaviour in infectious diseases models. Epidemics, 10:21–25, 2015. Challenges in Modelling Infectious DIsease Dynamics.

[56] Tiago P. Peixoto. Model selection and hypothesis testing for large-scale network models with overlapping groups. Phys. Rev. X, 5:011033, Mar 2015.

