## Supplementary Material for "Impact of the representation of contact data on the evaluation of interventions in infectious diseases simulations"

---

### S1 Data and methods

#### S1.1 Contact data

##### S1.1.1 Contact networks

We use empirical data describing contacts between individuals, collected using Radio Frequency Identification (RFID) devices that measure mutual proximity in a distributed fashion [1]. During each data collection period, individuals were asked to wear the RFID sensors on their chest, embedded in unobtrusive wearable badges. The sensors can exchange ultra-low power radio packets, and the power level is tuned so that a sensor can receive a packet emitted by another sensor only when the individuals wearing them face each other at close range (about 1 to 1.5 m). Moreover, the wearable sensors are tuned so that the face-to-face proximity of two individuals wearing them can be assessed over an interval of 20 seconds with a probability in excess of 99% [1]. Two individuals are thus said to be in contact if their badges exchange radio packets during a 20-second time window, and the contact event is considered interrupted if the badges do not exchange packets over a 20-second interval. We refer to [1] for more details. The data temporal resolution is thus of 20 seconds, and each data set consists in a temporal network giving for each time-window of 20 seconds the list of individuals who have been in contact during that time window. All such data sets are publicly available on the webpage `www.sociopatterns.org/datasets`.

For simplicity and better efficiency of the simulations, we aggregate the data on intervals of duration 15 minutes. We thus obtain on each $\delta t$ =15-minutes time-window a weighted network of interactions, where each node represents an individual, and each link between two nodes corresponds to the fact that these individuals have been in contact at least once during the time-window; the weight of each link gives the fraction of time during which the individuals have been in contact. This yields the DYN representation of the data.

We used contact data recorded in four distinct contexts: offices (OD), hospital (HD), a primary school (PS), and a high school (CP). In each case, we exclude days with incomplete recording, and hours without activity or with very low activity: this yields the days and time limits described in Table 3 of the main text. Figure S1 shows the corresponding periods and activity timelines, i.e., the total contact time in each time window of 15 minutes.

##### S1.1.2 Data representations

We define in the main text the various representations of the temporal contact data used in our simulations. We specify here that in this work, we have slightly modified the CMD (Contact Matrix of Distributions) representation with respect to its original definition [2]. Here indeed, the negative binomial distribution used to assign weight in the CMD representation fits only the non-zero weights found in the data. The correct densities of links are enforced by measuring and explicitly assigning the average daily number of contacts between each pair of categories.

For the CMD and CMB (Contact Matrix of Bimodal distributions) representations, we create 100 daily networks of contacts, used in a random order in each simulation using the corresponding representation.

#### S1.2 Numerical simulations

##### S1.2.1 Algorithm

Each simulation is initialized as follows:

- We choose at random an initial time between 0 and a maximum initial time. The maximum initial time for each setting is specified in Table S1.

- We initialize all the $N$ individuals in the susceptible compartment.

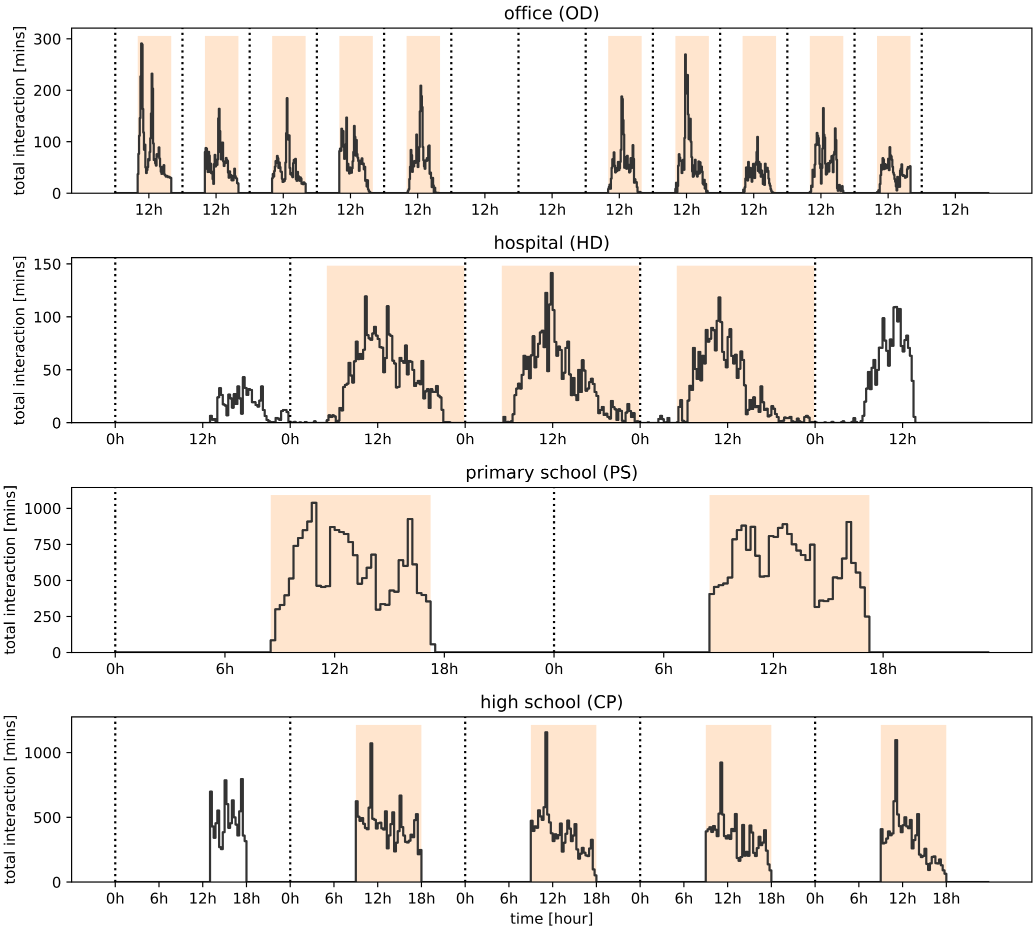

Figure S1: Timelines of contact activity for the empirical data. From top to bottom: OD, HD, PS, and CP. The line gives the value of the total time in contact of all individuals during each 15-minutes time window. Dashed vertical lines separate different days. Filled areas highlight the periods of activity considered in each setting.

Table S1: Maximum initial time for each setting

| | OD | HD | PS | CP |
|---|---|---|---|---|
| Max. initial time [days] | 14 | 3 | 7 | 7 |

- Given a value $i$ of the initial immunity, we choose at random $N_i = round(Ni)$ individuals to start in the recovered compartment.
- Given a vaccine coverage $v$, we choose at random $N_v = round(Nv)$ individuals to have the parameters $\sigma, p_c, r_\beta$ associated with the vaccine.
- Among the remaining susceptible and non-vaccinated population, we choose one individual at random to be in the exposed compartment at the initial time.
- In the case of regular testing (RT) and reactive screening (rT) with adherence $\alpha$, we choose at random $N_\alpha = round(N(1-v)\alpha)$ individuals among the non-vaccinated to accept to perform the tests. In the case of telework, we define random groups to alternate.
- In the case of CMB and CMD, we shuffle the collection of pre-generated random graphs.

Simulations are performed using discrete time steps of $\delta t = 15$ minutes. During each time step we perform the following procedures:

- We evaluate new infections from infectious individuals towards their neighbors at that time step (as informed by the data) who are not in quarantine or telework (see S1.2.2).
- We perform the actions associated with the active NPI at that time, if any. Possible actions include testing for individuals who just entered the symptomatic compartment, scheduled regular or reactive testing, change in the quarantine status for a individual, etc (see S1.2.3).
- We check if the infection status of each exposed or infected individual changes at that time (see S1.2.4).
- We check whether to introduce a new exposed individual (see S1.2.5)

Depending on the purpose of the simulations, they can be run until a specific condition is satisfied (e.g., when no new cases are introduced, until no infectious remains), or for a fixed duration. For each run, we record the time of infection for each infected individual (and who infected them). We also save the number of days of quarantine.

To evaluate NPIs, we compute a bootstrapped mean and median epidemic size, the cost of the protocol as the average number of days of quarantine per individual, and efficacy measured as the relative reduction of the median size compared with symptomatic testing in the same representation and conditions (see S1.4).

##### S1.2.2 Evaluation of new infections

At each time step of activity in a given setting (see Table 3 of the main text), we evaluate for each infectious individual the possible new infections towards susceptible neighbors. The list of neighbors is given by the weighted graph corresponding to that time. For the DYN representation, the network changes at every time step. For dHET (daily heterogeneous networks), CMD and CMB it changes every day. For HET (heterogeneous networks), CM (Contact Matrix) and FULL, the network is constant over time.

For DYN, the weight is directly taken from the weighted network. For all the others, the weight is multiplied by $\delta t / T_{day}$, where $T_{day} = t_f - t_i$ is the total daily time of activity in the setting (see Table

3 of the main text) and $\delta t = 15$ minutes is the simulation time step. Naturally, we exclude neighbours who are in quarantine from the evaluation of possible new infections.

For a susceptible neighbour (with susceptibility $\sigma$) of an infectious individual in the compartment $X \in I_p, I_{sc}, I_c$, with relative infectiousness $r_\beta^X$, the probability to become infected after being in contact for a time $w(t)$ at time $t$ is given by:

$$p = 1 - \exp\left(-\sigma r_\beta^X \beta w(t)\right). \tag{S1}$$

##### S1.2.3 Intervention details

Each individual, independently from their infection status, has a quarantine status, which lasts a determined time depending on the reason which triggered the change in status. When an individual is quarantined, their disease keeps evolving, but they will not interact with other individuals until their quarantine ends.

Table S2: Parameters for the considered interventions, taken from [3]

| parameter | | value |
|---|---|---|
| probability of detection | $p_D$ | |
| (adults and adolescents) | | 0.5 |
| (children) | | 0.3 |
| sensitivity $I_p$ | $\theta_p$ | 0.5 |
| sensitivity $I_c$ | $\theta_c$ | 0.8 |
| sensitivity $I_{sc}$ | $\theta_{sc}$ | 0.7 |
| quarantine period | $\Delta_Q$ | 7 days |
| reactive period | $\Delta_r$ | 1 day |
| reactive period | $\Delta_r^2$ | 4 day |
| turnaround time | $\Delta_w$ | 15 mins |

Antigen tests modeled as shown in Table S2 are used in NPIs evaluation. For sensitivity, we compared the performance of NPIs using a PCR-test model in section S2.5.1.

**Testing timing.** Symptomatic tests are performed outside the hours of activities in the first possible moment after an individual enters the clinical compartment $I_c$. Regular testing is performed simultaneously for all compliant individuals on Tuesday at the initial hour of activities. In the case of regular tests twice per week, they are performed on Tuesdays and Thursdays.

Reactive testing in schools is performed simultaneously for all the compliant individuals, $\Delta_Q = 1$ day after the positive case is detected, and again $\Delta_{Q2} = 4$ days after.

**Teacher replacement.** In the primary school setting, if a teacher tests positive, the teacher is replaced by a new susceptible individual. After completing the quarantine, the teacher tests again: If negative, the teacher comes back to school (possibly recovered, but not necessarily as the tests can yield false negatives); if positive, the teacher remains in quarantine for a second time.

##### S1.2.4 Compartment evolution and time distribution in the epidemic model

While infections and tests occur on a time-step basis, the evolution of an infection within a host is event-driven. The duration in a compartment is defined right after a individual enters a compartment: it is drawn from a Gamma distribution using the parameters given in Table S3.

Table S3: Parameters for the Gamma distribution used in our simulations for the compartmental model. Taken from [3]

| SEIR parameter | shape $k$ | scale $\theta$ |
|---|---|---|
| $\tau_E$ | 3 | 4/3 days |
| $\tau_p$ | 1 | 1.8 days |
| $\tau_I$ | 6 | 5/6 days |

###### S1.2.5 Weekly introductions

For each block of $n_w$ weeks, we pick with a uniform probability a random time $t_w$ out of activity hours to convert a susceptible individual into an exposed one. The probability of being chosen is equal for all individuals, regardless of their age or vaccination status.

Introductions are considered when evaluating NPIs. We used $n_w = 2$ for most of our analysis, and $n_w = 1$ for sensitivity in Section S2.5.2.

##### S1.3 Infection rate calibration

We perform 4000 independent runs, varying the value of the rate of transmission $\beta$, starting with one initial seed, and running each simulation until the recovery of the initial infectious seed. We use as definition for the basic reproduction number $R_0$ the average number of first generation infections per seed, and fit the results using the function

$$R_0^{fit}(\beta) = n_f(1 - exp(-\beta\tau_f)) \ . \tag{S2}$$

The interval of values of $\beta$ considered for the calibration is specific to each setting and representation, defined after a pre-calibration used to estimate the values of $\beta$ such that $R_0$ lies approximately between $R_0 = 0.75$ and $R_0 = 4$. We use 30 values of $\beta$ logarithmically distributed (figure S2).

Equation (S2) can be inverted to obtain the value of $\beta$ such that the reproductive number is $R_0$ under representation $rep$, denoted as $\beta_{rep}(R_0)$. To compare representations we can define the scaling factors between representations $p_{rep1|rep0}(R_0) = \beta_{rep1}(R_0)/\beta_{rep0}(R_0)$. We show the scaling factors in different settings in figure S2. Dense representations (e.g. CM) are more sensitive to variation on the rate of transmission with respect to sparse ones (e.g. HET).

##### S1.4 Bootstrapping and confidence intervals

All presented results are obtained bootstrapping the statistics from different runs, as follows.

Given a collection of $n$ independent observations $X = \{x_i\}_{i=1,...,n}$ drawn from an unknown distribution, we are interested in estimating the value and confidence interval of a function $f(X)$ (for instance, the median, the mean, or the proportion of observations satisfying a condition). Bootstrap generates $m$ re-samples with replacement of size $n$ from the collection $X$, evaluating the function $f$ on each re-sample to create a distribution of estimators $F = \{f_j\}_{j=1,...,m}$. The estimated value of the function is given by $mean(F)$, and the confidence intervals are given by excluding the $1 - \alpha/2$ left and right percentiles, with $n = 2000$, $m = 4000$ and $\alpha = 5\%$ [4].

When comparing the reduction $r(X|Y)$ of the epidemic size under the intervention $P_X$ with respect to the intervention $P_Y$, where we have $n$ observations of the epidemic size for each intervention, respectively $X = \{x_i\}$ and $Y = \{y_i\}$, we generate $m$ re-samples with replacement $X'_m$ and $Y'_m$ of size $n$ from both collections, and we compute the reduction estimated for each pair of re-samples $R(X|Y) = 1 - median(X'_m)/median(Y'_m)$.

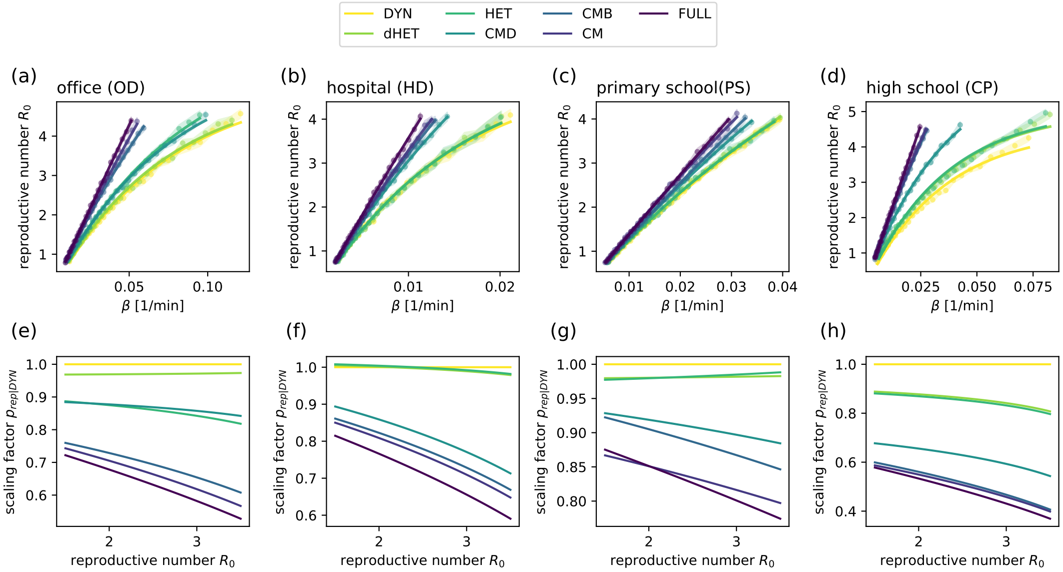

Figure S2: Calibration Rate of transmission on the different settings. (a,b,c,d) Reproductive number $R_0$ versus rate of transmission $\beta$ for different representations. Dots are the result of numerical simulation, and the continuous lines correspond to the fits to Eq. (S2). Shaded areas correspond to the estimated error, obtained as a bootstrapped CI. (e,f,g,h) Scaling factors for different representations compared with dynamical networks (DYN), vs. $R_0$. The panels correspond to (a,e) offices, (b,f) hospital, (c,g) primary school, (d,h) high school.

#### S2 Supplemental Results

##### S2.1 Comparing scales of temporal aggregation

Considering daily contact graphs (dHET) yields results very close to the ones obtained using dynamical networks (DYN), compared with the use of heterogeneous networks aggregating all days (HET). We explain this observation by the fact that using a temporal resolution much finer than the time-scales of the infection process has little impact [5]. An alternative hypothesis would be that using a different graph each day modulates the number of contacts, as we observe an important variation in activity across days (see figure S1). To test this, we run simulations using daily contact matrices (dCM) that respect the daily modulation of activity. However, we observe almost no difference between dCM and CM (figure S3).

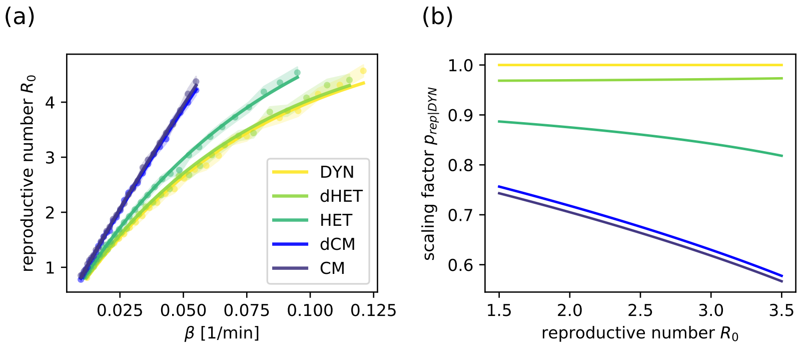

Figure S3: Effect of taking into account daily changes. (a) Reproductive number $R_0$ versus rate of transmission $\beta$ in the office dataset, comparing the individual based representations DYN, dHET, and HET on one hand, and the category-based representations CM and dCM (daily contact matrix) on the another hand. Dots are the result of numerical simulations, and the continuous lines correspond to the fit of Eq. (S2). Shaded areas correspond to the estimated error, obtained as a bootstrapped CI. (b) Scaling factors for the same representations compared with DYN representation.

##### S2.2 Cascading and effect of immunity

Figure 2 in the main text shows the distribution of reproductive numbers and the cascading effect in the office data set OD. Here we present the same analysis including $R_0 = 1.5$ (Figure S4) and the analysis on the other three data sets (Figure S5 for HD, Figure S6 for PS, and Figure S7 for CP).

Table S4 moreover gives the parameters of the negative binomial fits to the distributions of reproductive numbers.

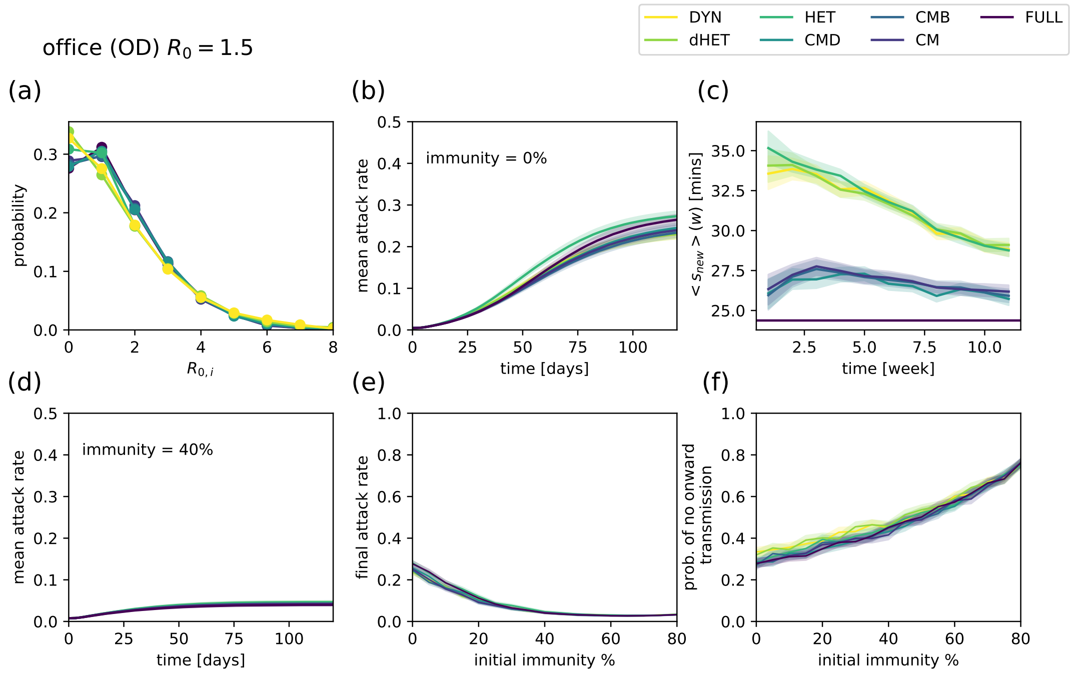

Figure S4: Spreading dynamics on different representations of the office data set, for $R_0 = 1.5$, starting from a single initial exposed seed (results for $R_0 = 3.0$ are shown in figure 2 of the main text). (a) Distribution of the number of secondary infections produced by the initial seed. (b) Temporal evolution of the mean attack rate from one single exposed individual in a otherwise susceptible population. (c) Average strength of newly infected individuals infected in a given week vs. time. In panels (a,b,c), initial immunity is set to zero. (d) Temporal evolution of the mean attack rate, starting from one single exposed individual in a population with an initial immunity of 40%. (e) Mean final attack rate vs. initial immunity for different representations (f) Probability of no onward transmission (i.e. the initial seed does not infect any other individual) as a function of the initial immunity. Shaded areas correspond to the estimated error, obtained as a bootstrapped CI.

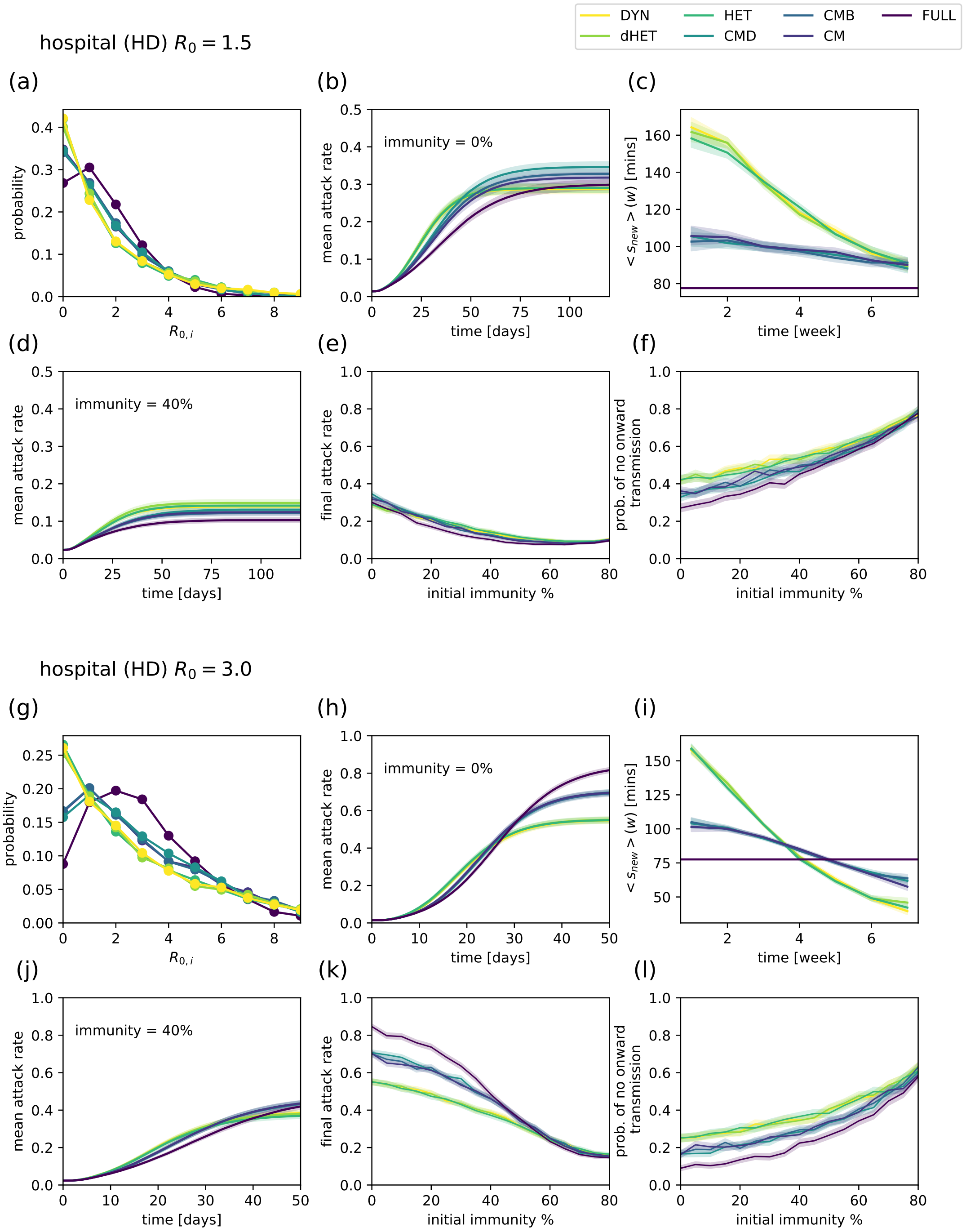

Figure S5: Spreading dynamics on different representations of the hospital data set, for $R_0 = 1.5$ and $R_0 = 3.0$, starting from a single initial exposed seed. (a,g) Distribution of the number of secondary infections produced by the initial seed. (b,h) Temporal evolution of the mean attack rate from one single exposed individual in a otherwise susceptible population. (c,i) Average strength of newly infected individuals infected in a given week vs. time. In panels (a,b,c,g,h,i), initial immunity is set to zero. (d,j) Temporal evolution of the mean attack rate, starting from one single exposed individual in a population with an initial immunity of 40%. (e,k) Mean final attack rate vs. initial immunity for different representations (f,l) Probability of no onward transmission as a function of the initial immunity. Shaded areas correspond to the estimated error, obtained as a bootstrapped CI.

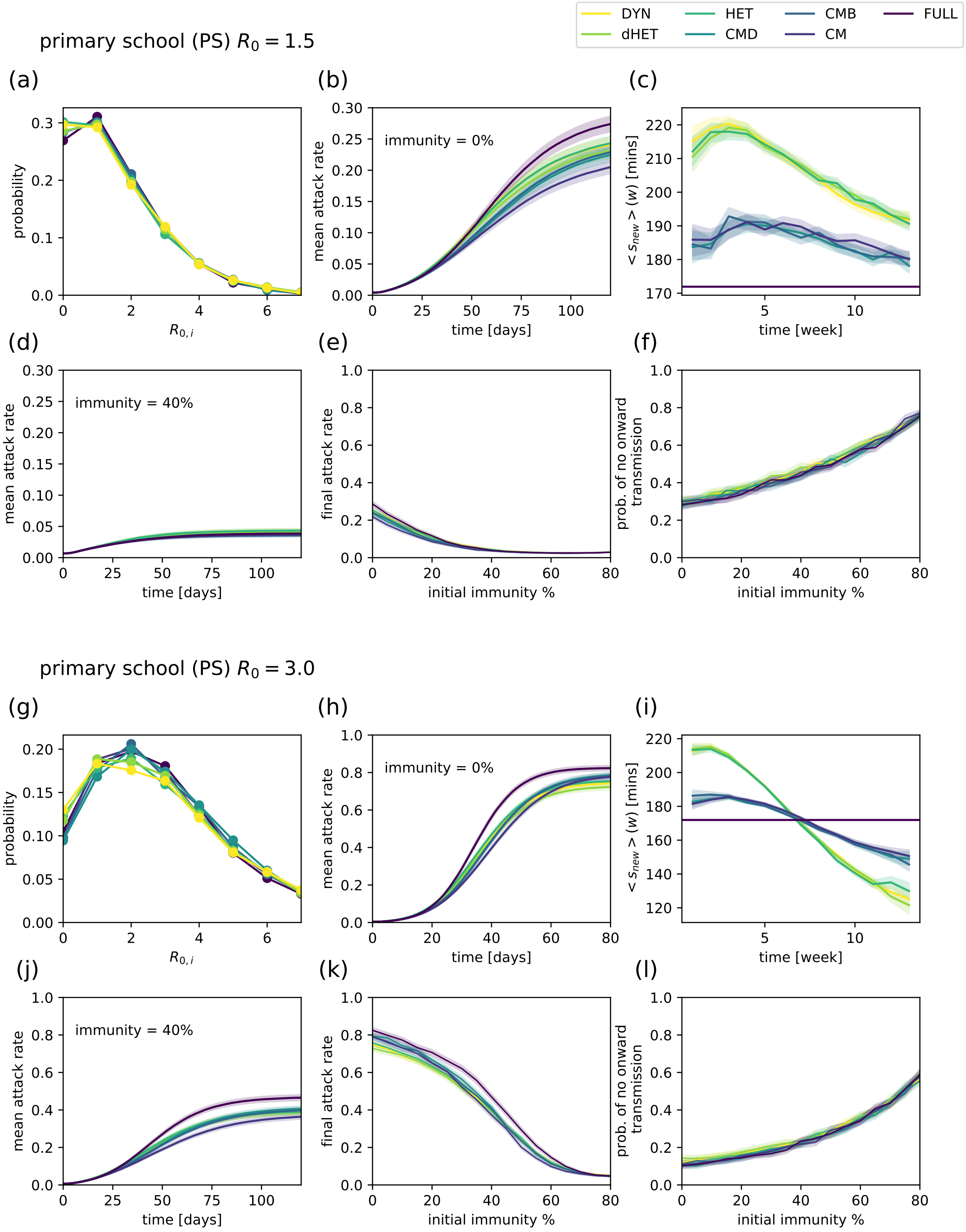

Figure S6: Spreading dynamics on different representations of the primary school data set, for $R_0 = 1.5$ and $R_0 = 3.0$, starting from a single initial exposed seed. (a,g) Distribution of the number of secondary infections produced by the initial seed. (b,h) Temporal evolution of the mean attack rate from one single exposed individual in a otherwise susceptible population. (c,i) Average strength of newly infected individuals infected in a given week vs. time. In panels (a,b,c,g,h,i), initial immunity is set to zero. (d,j) Temporal evolution of the mean attack rate, starting from one single exposed individual in a population with an initial immunity of 40%. (e,k) Mean final attack rate vs. initial immunity for different representations (f,l) Probability of no onward transmission as a function of the initial immunity. Shaded areas correspond to the estimated error, obtained as a bootstrapped CI.

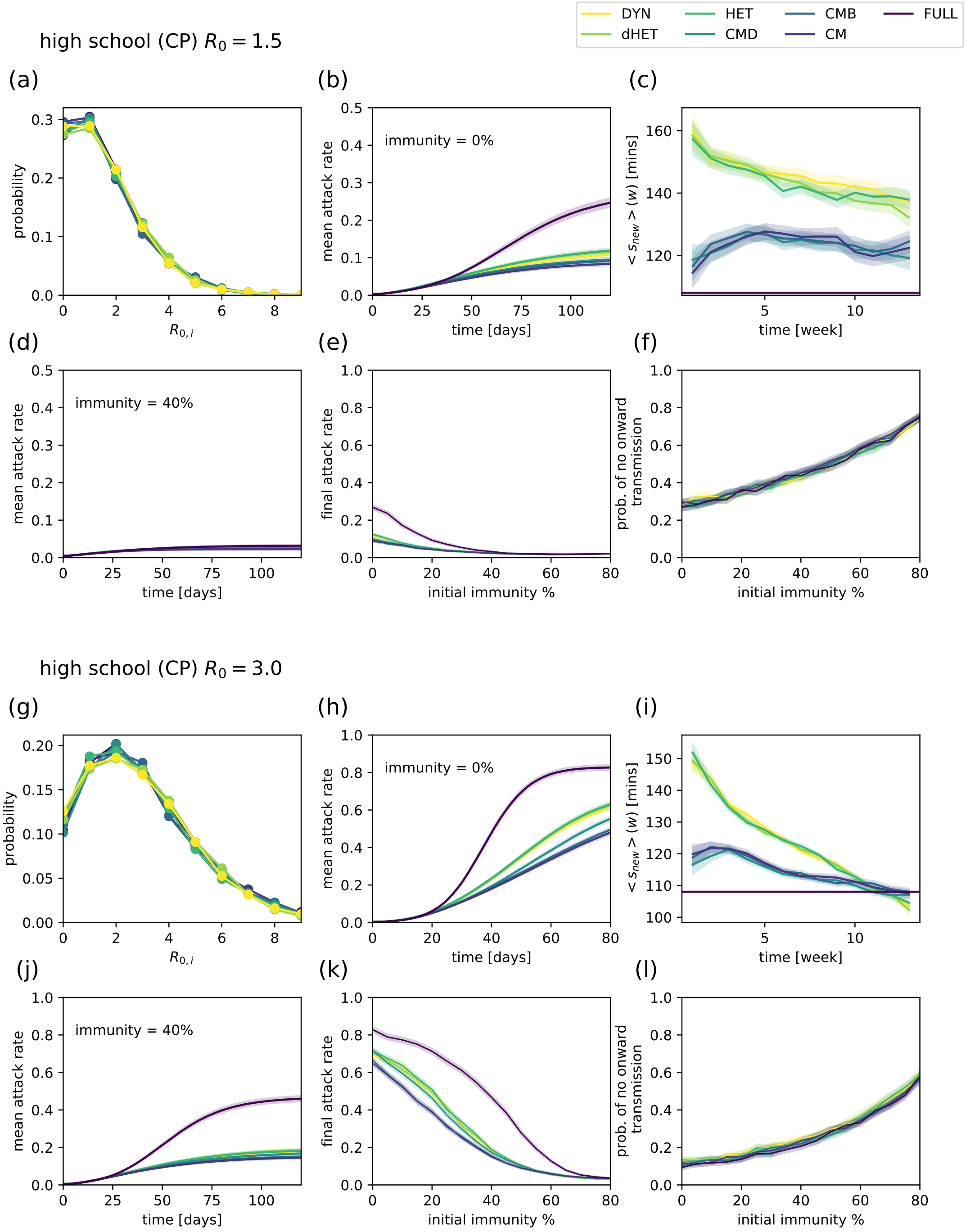

Figure S7: Spreading dynamics on different representations of the high school data set, for $R_0 = 1.5$ and $R_0 = 3.0$, starting from a single initial exposed seed. (a,g) Distribution of the number of secondary infections produced by the initial seed. (b,h) Temporal evolution of the mean attack rate from one single exposed individual in a otherwise susceptible population. (c,i) Average strength of newly infected individuals infected in a given week vs. time. In panels (a,b,c,g,h,i), initial immunity is set to zero. (d,j) Temporal evolution of the mean attack rate, starting from one single exposed individual in a population with an initial immunity of 40%. (e,k) Mean final attack rate vs. initial immunity for different representations (f,l) Probability of no onward transmission as a function of the initial immunity. Shaded areas correspond to the estimated error, obtained as a bootstrapped CI.

Table S4: Over-dispersion parameters of the distribution of the number of secondary infections produced by the initial seed, obtained by fitting the distribution to a negative binomial distribution: dispersion parameter $k$ (inverse of the number of trials $r = 1/k$), and probability $p$, for different settings and representations, with $R_0 = 1.5$ and $R_0 = 3.0$.

| | $R_0$ | 1.5 | | 3.0 | | | $R_0$ | 1.5 | | 3.0 | |
|---|---|---|---|---|---|---|---|---|---|---|---|
| setting | rep. | $k$ | $p$ | $k$ | $p$ | setting | rep. | $k$ | $p$ | $k$ | $p$ |
| OD | DYN | 0.51 | 0.56 | 0.47 | 0.42 | PS | DYN | 0.33 | 0.67 | 0.33 | 0.50 |
| | dHET | 0.56 | 0.54 | 0.49 | 0.41 | | dHET | 0.33 | 0.66 | 0.28 | 0.54 |
| | HET | 0.40 | 0.63 | 0.33 | 0.51 | | HET | 0.35 | 0.66 | 0.29 | 0.53 |
| | CMD | 0.25 | 0.72 | 0.18 | 0.65 | | CMD | 0.26 | 0.72 | 0.18 | 0.64 |
| | CMB | 0.27 | 0.71 | 0.25 | 0.57 | | CMB | 0.27 | 0.71 | 0.21 | 0.61 |
| | CM | 0.25 | 0.73 | 0.27 | 0.56 | | CM | 0.26 | 0.72 | 0.23 | 0.59 |
| | FULL | 0.22 | 0.75 | 0.23 | 0.59 | | FULL | 0.23 | 0.74 | 0.23 | 0.60 |
| HD | DYN | 1.14 | 0.37 | 0.96 | 0.26 | CP | DYN | 0.26 | 0.72 | 0.25 | 0.58 |
| | dHET | 1.05 | 0.38 | 0.94 | 0.26 | | dHET | 0.25 | 0.72 | 0.21 | 0.62 |
| | HET | 1.18 | 0.36 | 0.98 | 0.26 | | HET | 0.28 | 0.70 | 0.24 | 0.59 |
| | CMD | 0.57 | 0.54 | 0.48 | 0.41 | | CMD | 0.24 | 0.74 | 0.20 | 0.63 |
| | CMB | 0.57 | 0.54 | 0.52 | 0.39 | | CMB | 0.32 | 0.67 | 0.24 | 0.58 |
| | CM | 0.58 | 0.54 | 0.51 | 0.39 | | CM | 0.29 | 0.70 | 0.24 | 0.58 |
| | FULL | 0.18 | 0.79 | 0.18 | 0.65 | | FULL | 0.24 | 0.74 | 0.23 | 0.59 |

#### S2.3 Evaluation of NPIs

##### S2.3.1 NPIs evaluation for different settings and data representations

Figure 4 in the main text shows the evaluation of a selection of NPIs for the office and primary school data. Here we expand these results, with more NPIs, $R_0 = 1.5$ and $R_0 = 3$, and the four settings.

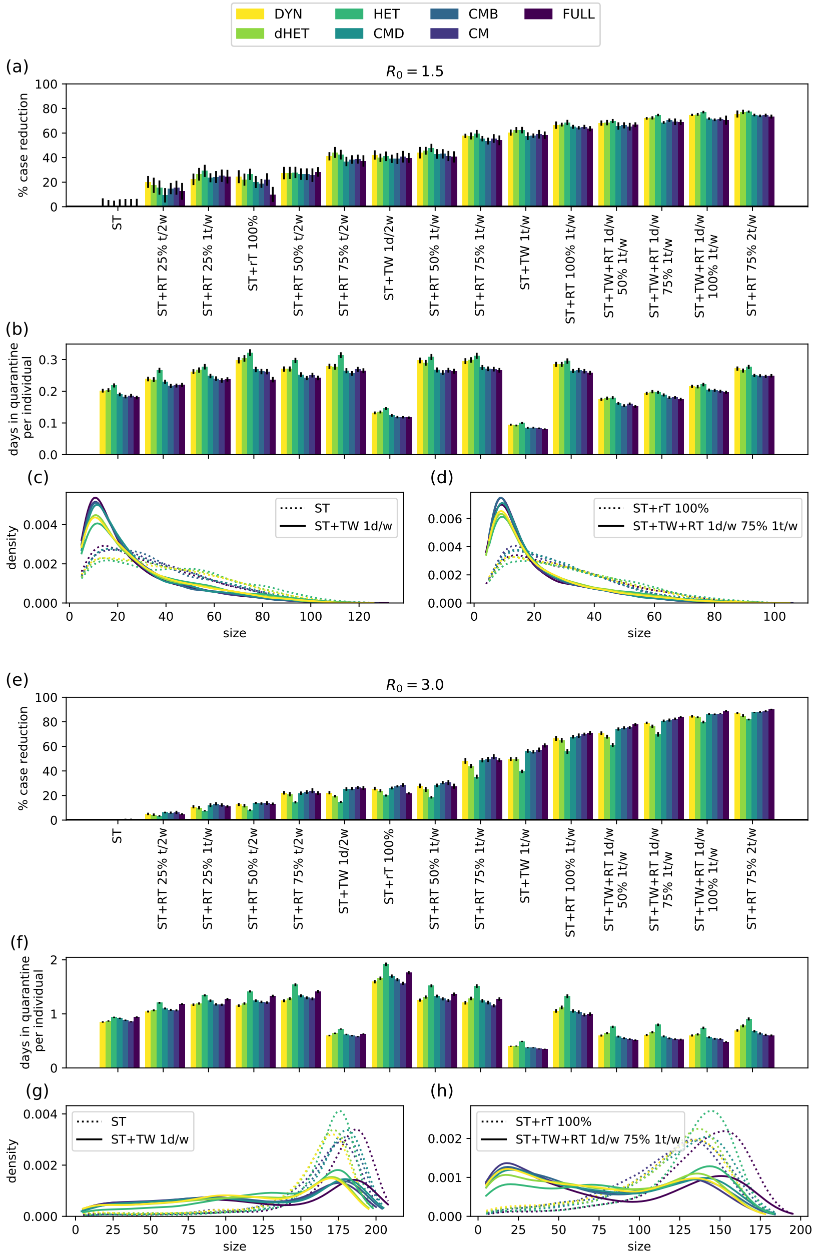

Figure S8: Evaluation of NPIs in offices (OD) for $R_0 = 1.5$ and $R_0 = 3.0$, and simulations performed using different data representations. (a,e) Efficacy of NPIs sorted by increasing order of efficacy in the DYN representation. Efficacy is defined as the relative reduction in median size compared with symptomatic testing (ST) alone, after a period of 60 days. (b,f) Average number of days in quarantine per individual under different protocols. (c-d,g-h) Epidemic size distributions for several protocols.

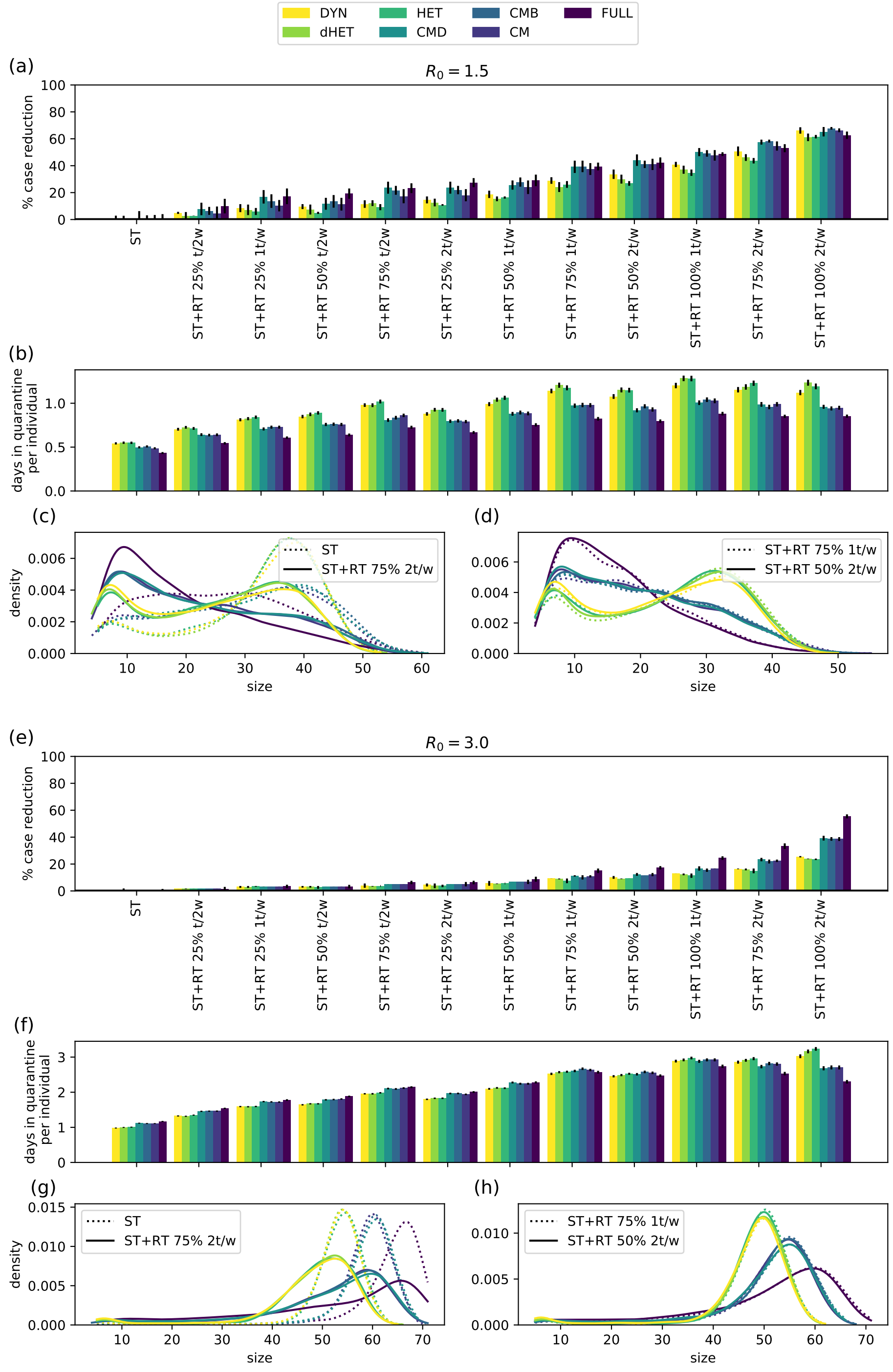

Figure S9: Evaluation of several NPIs in the hospital dataset (HD) for $R_0 = 1.5$ and $R_0 = 3.0$, and simulations performed using different data representations. (a,e) Efficacy of NPIs sorted by efficacy increasing order. Efficacy is defined as the relative reduction in median size compared with symptomatic testing (ST) alone, after a period of 60 days. (b,f) Average number of days in quarantine per individual under different protocols. (c,d,g,h) Epidemic size distributions for different protocols. The protocols ST+RT with respectively $\alpha = 75\%$, frequency once per week and $\alpha = 50\%$, frequency twice per week have not only similar efficacy, but also the shapes of the distributions of epidemic sizes are very similar: this similarity holds across diverse representations.

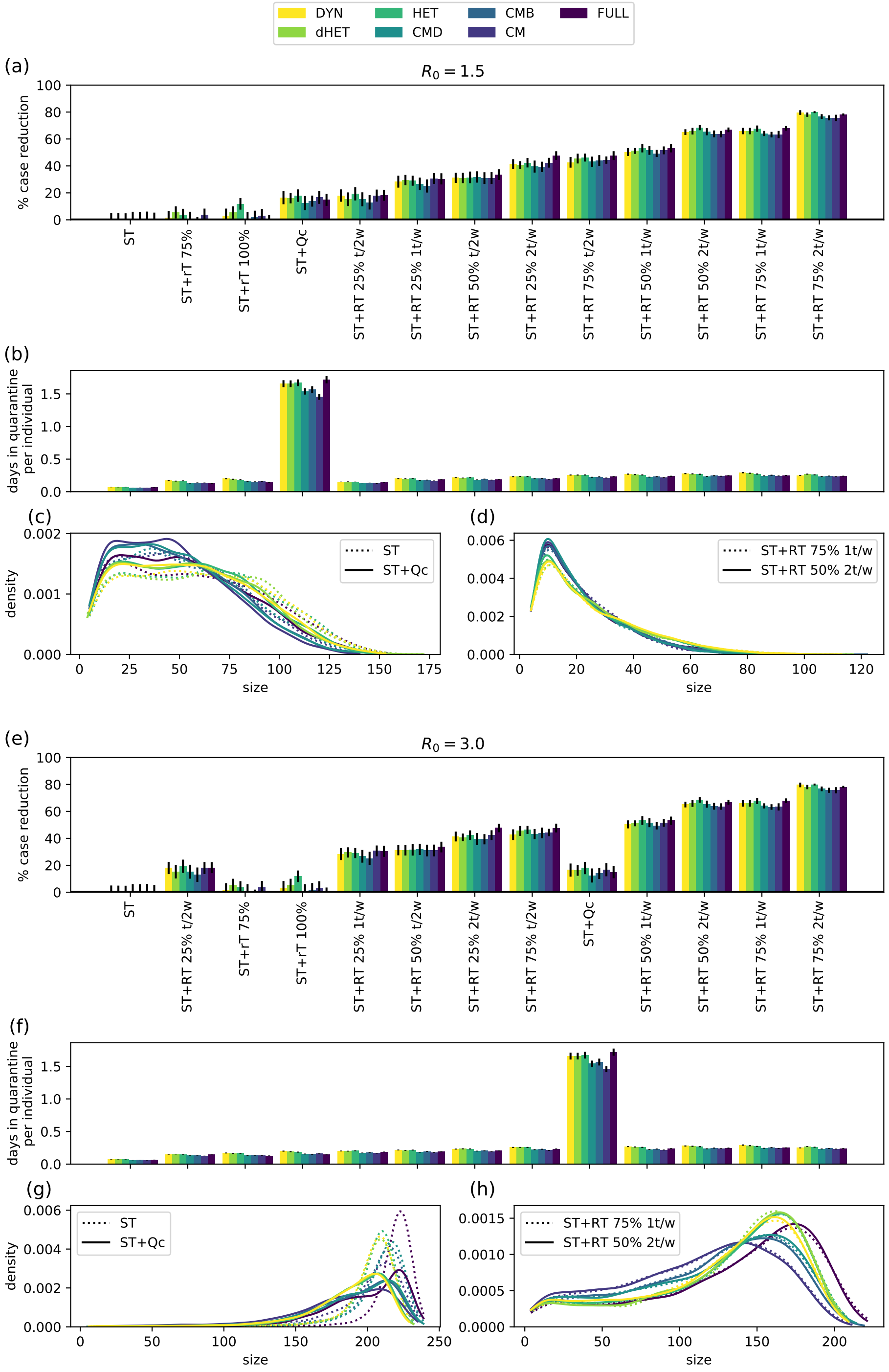

Figure S10: Evaluation of several NPIs in the primary school dataset (PS) for $R_0 = 1.5$ and $R_0 = 3.0$, and simulations performed using different data representations. (a,e) Efficacy of NPIs sorted by efficacy increasing order. Efficacy is defined as the relative reduction in median size compared with symptomatic testing (ST) alone, after a period of 60 days. (b,f) Average number of days in quarantine per individual under different protocols. (c-d,g-h) Epidemic size distributions for different protocols. The protocols ST+RT with respectively $\alpha = 75\%$, frequency once per week and $\alpha = 50\%$, frequency twice per week have not only similar efficacy, but also the shapes of the distributions of epidemic sizes are very similar: this similarity holds across diverse representations.

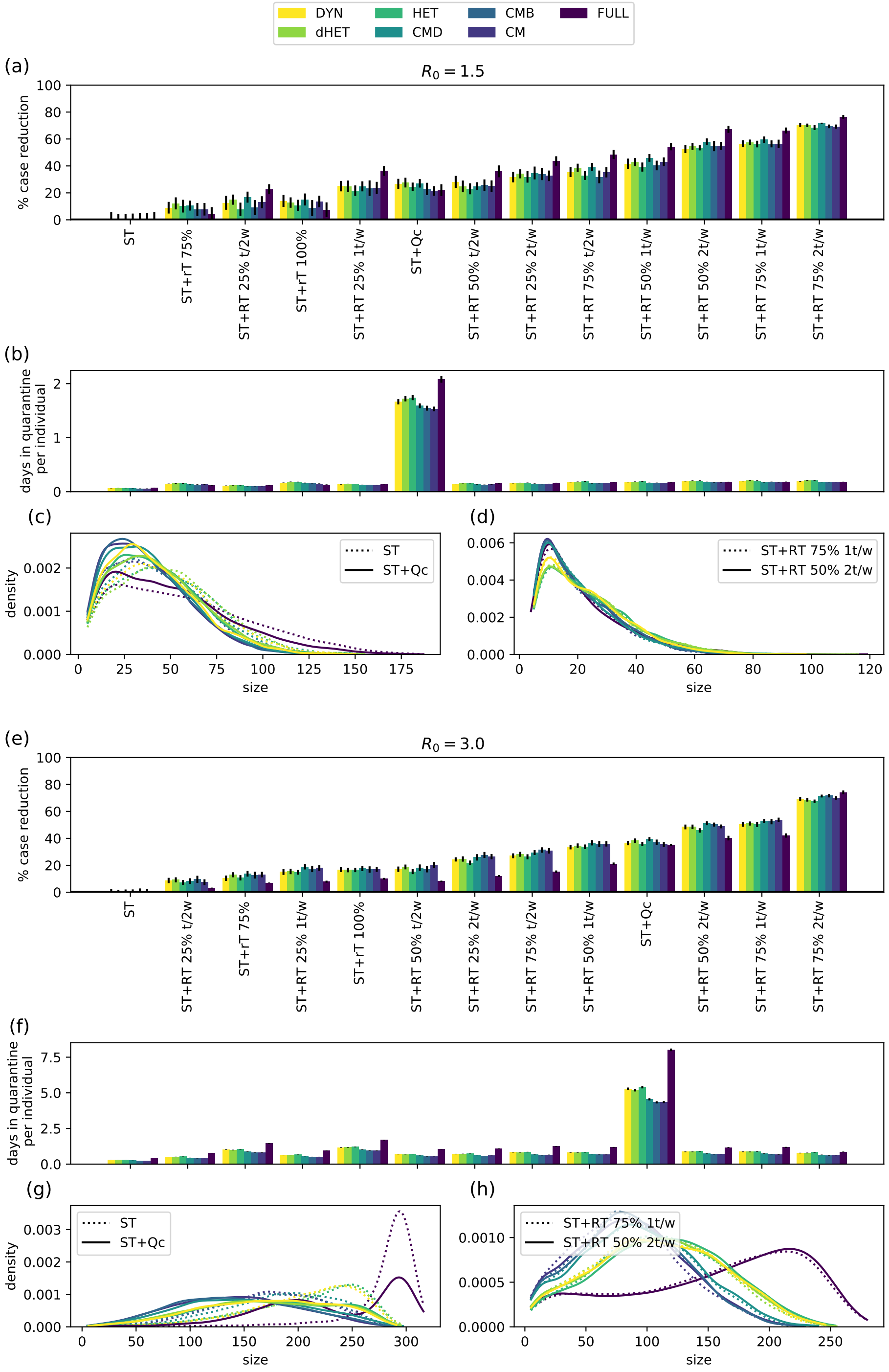

Figure S11: Evaluation of several NPIs in the high school dataset (CP) for $R_0 = 1.5$ and $R_0 = 3.0$, and simulations performed using different data representations. (a,e) Efficacy of NPIs sorted by efficacy increasing order. Efficacy is defined as the relative reduction in median size compared with symptomatic testing (ST) alone, after a period of 60 days. (b,f) Average number of days in quarantine per individual under different protocols. (c-d,g-h) Epidemic size distributions for different protocols. The protocols ST+RT with respectively $\alpha = 75\%$, frequency once per week and $\alpha = 50\%$, frequency twice per week have not only similar efficacy, but also the shapes of the distributions of epidemic sizes are very similar: this similarity holds across diverse representations.

##### S2.3.2 NPI efficacy vs. basic reproductive number

In the main text, we discuss how NPIs have a maximum in their relative efficacy at a certain value of the reproductive number, which is specific for each setting and NPI. Figure S12 shows this behaviour for four selected NPIs in the office setting.

The differences in relative efficacy measured using different representations are small at small values of $R_0$. They become larger at intermediate values of $R_0$. For very large values of $R_0$ in which all NPIs have a very small efficacy, differences decrease again (here, seen only for ST).

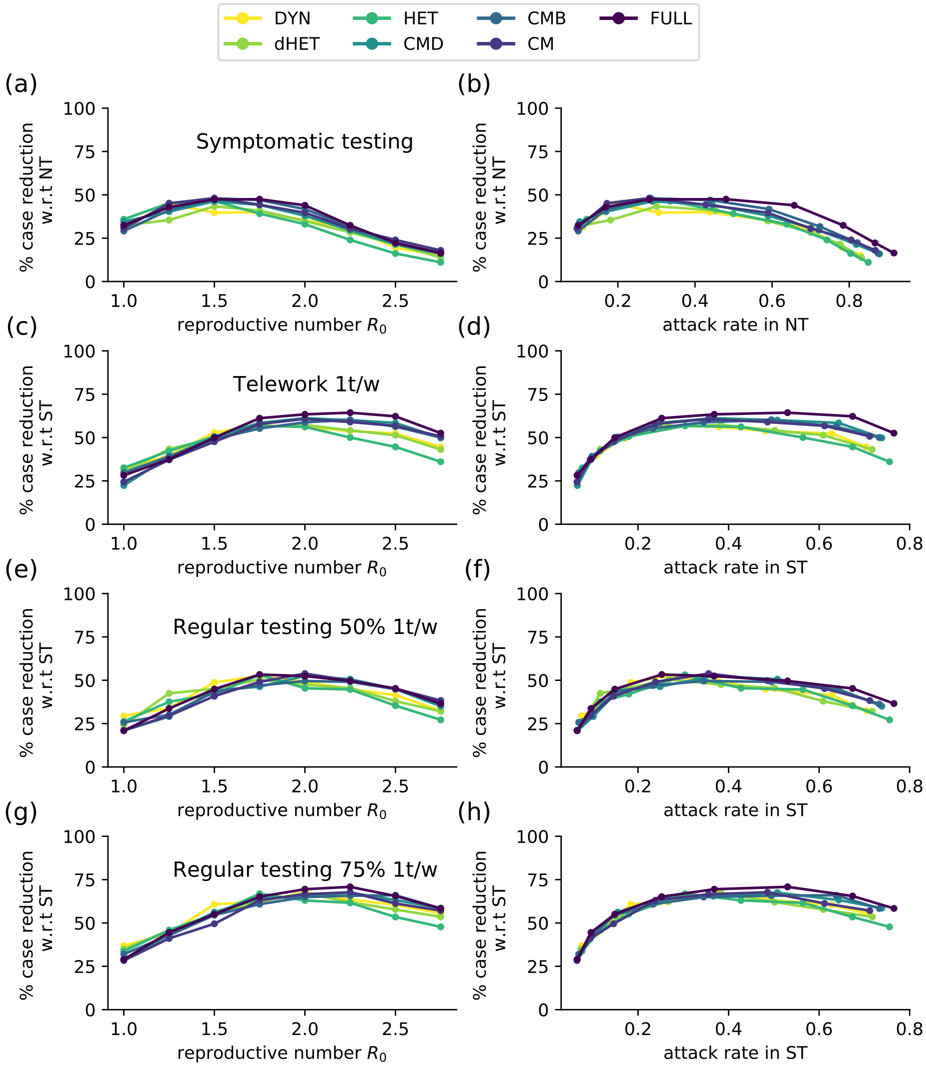

Figure S12: In the left panels, we show the relative reduction in epidemic size for selected NPIs as the reproductive number increases between $R_0 = 1.0$ and $R_0 = 3.0$, in the office data set. In the right panels, we compare this reduction with the attack rate in the baseline scenario (ST). The efficacy of NPIs are measured as relative with respect to ST, except for ST itself which is compared against NT.

##### S2.3.3 Adherence vs. frequency in regular testing

We show here additional results on an increase in the number of tests performed in RT protocols, through an increase in adherence or in frequency. Figure S13 extends the results shown in Figure 5c,d: the average size reduction per test is shown for ST+RT with frequency once per week and adherence 25% (left column) or 50% (right column): it is given by $r_0/n_T^0$ where $r_0$ is the size reduction and $n_T^0$ the number of tests for this protocol. The new size reduction is noted $r_\alpha$ when increasing the adherence $\alpha$ and $r_f$ when increasing the frequency. The corresponding increases in the numbers of tests are denoted $\Delta n_T^\alpha$ and $\Delta n_T^f$ [1], and Figure S13 shows the additional size reduction per test obtained by doubling either the adherence $((r_\alpha - r_0)/\Delta n_T^\alpha)$ or the frequency $((r_f - r_0)/\Delta n_T^f)$. The additional size reduction per test is lower in both cases, with the exception of the hospital setting when increasing the adherence. Moreover, the additional efficacy per test is larger when doubling the adherence than when doubling the frequency, for all settings and all data representations.

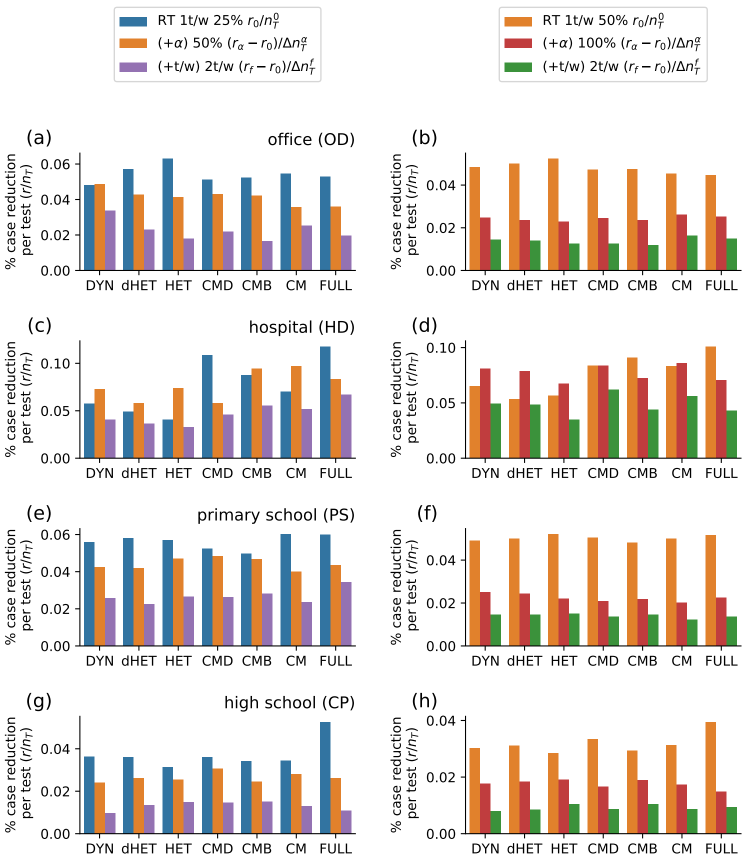

Figure S13: Effect of improving adherence or frequency, for $R_0 = 1.5$. Each panel shows (i) the average size reduction (w.r.t. ST) per test, for RT at frequency once per week and adherence 25% (left column) or 50% (right column): the additional size reduction per additional test (ii) when doubling the adherence, (iii) when doubling the frequency,

[1] Note that $\Delta n_T^\alpha \approx \Delta n_T^f$ but the two quantities are not strictly equal because the number of tests due to the ST protocol are slightly different (a protocol with larger efficacy reduces the number of symptomatic cases, this leads to a change of at most a few percents in the number of tests).

##### S2.3.4 Effect of initial immunity

Setting an initial portion of the population in the recovered state decreases the attack rate and the cost for all protocols, but does not affect the qualitative results. We show this here for the office setting.

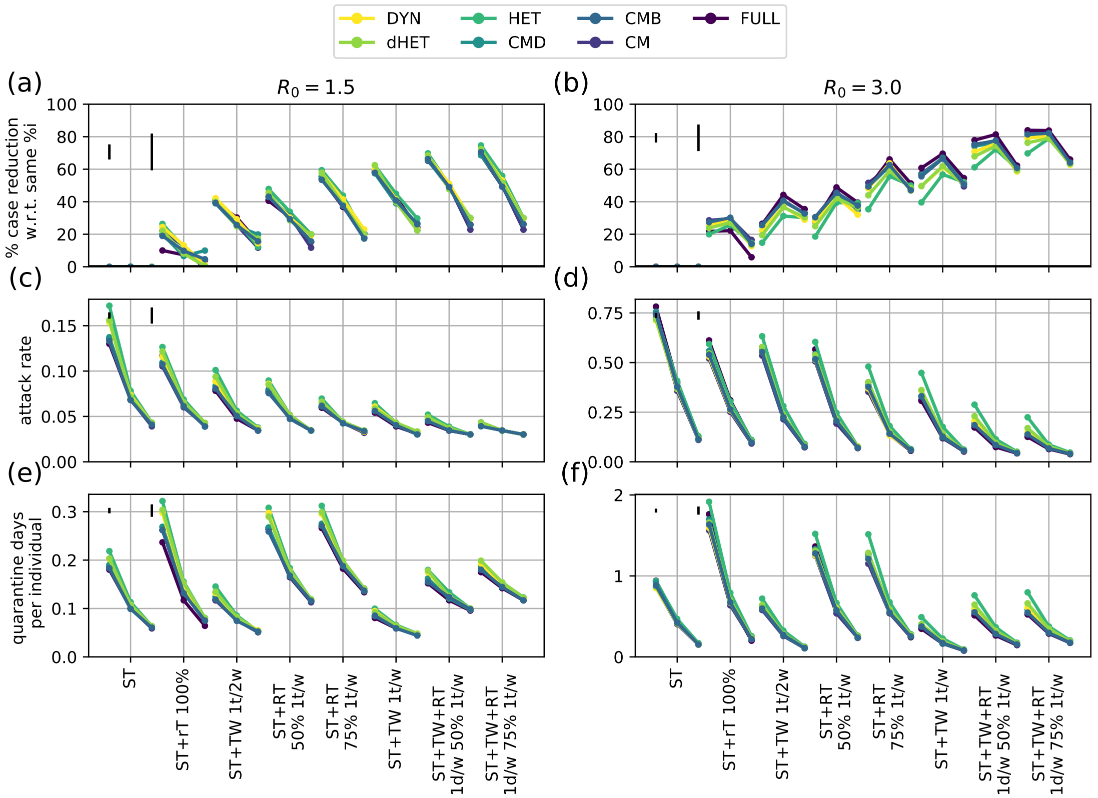

Figure S14: Effect of initial immunity in the office setting, considering reproductive numbers $R_0 = 1.5$ (left) and $R_0 = 3.0$ (right). For each protocol and representation, points represent an immunity of 0%, 25%, and 50%, from left to right. (a,b) Relative reduction with respect to ST, with the same level of immunity; (c,d) Attack rate at 60 days; (e,f) Days in quarantine per individual. Black vertical lines in the upper-left corner of each panel represent the median and maximum length of error bars among the data points.

#### S2.4 Combined effect of NPIs and vaccination

While NPIs are the only response to an emerging infectious disease as long as drugs and vaccines are not available, the context changes when a vaccine has been developed, as has been the case in the COVID-19 pandemic. It is thus also necessary to take into account how vaccination impacts the spread and the efficacy and cost of NPIs.

We explore this point in Figure S15 for the office data, with $R_0 = 1.5$ and $R_0 = 3$, and four values of the vaccination coverage (0%, 25%, 50%, and 75% of individuals, randomly selected to be vaccinated at the beginning of each simulation). Vaccination reduces the final epidemic size even in the absence of NPIs or for the basic ST protocol, and decreases the costs in terms of quarantines. The reduction in epidemic size with respect to the case of ST with no vaccination improves thus in all cases (and for all representations, Figure S15c,d,e,f).

The picture is more complex when taking as baseline the ST with the same vaccination coverage.

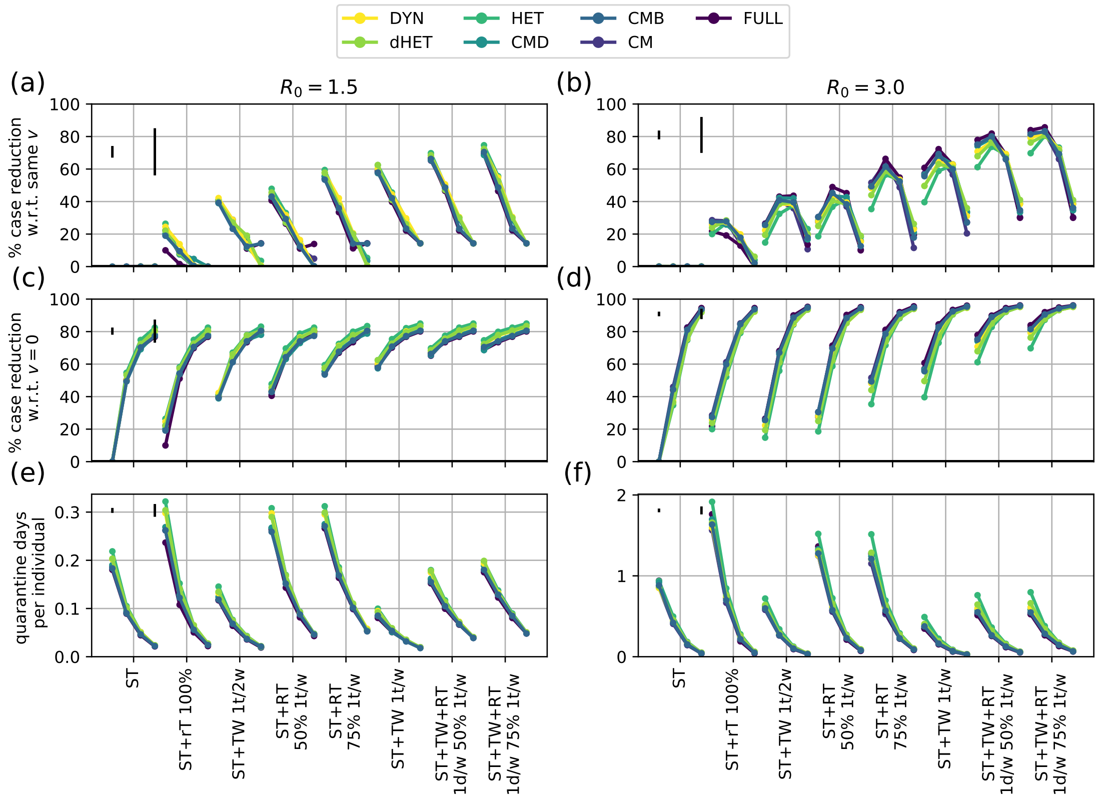

Figure S15: Effect of vaccine coverage for different NPIs in offices, considering reproductive numbers $R_0 = 1.5$ (left) and $R_0 = 3$ (right) and various data representations, in the office setting. For each protocol, the points represent a coverage of 0%, 25%, 50%, and 75% from left to right. (a-b) Relative reduction in median size with respect to ST with the same level of vaccine coverage (c-d) Relative reduction in median size with respect to ST with 0% vaccine coverage. (e-f) Days in quarantine per individual for different NPIs. Black vertical lines in the upper-left corner of each panel represent the median and maximum length of error bars among the data points.

For $R_0 = 1.5$, increasing vaccination leads to a decrease in the efficacy (Figure S15a), while it leads to a non-monotonic curve for $R_0 = 3$ (Figure S15b, see also [3]). This is due to the non-monotonic behaviour of the efficacy with respect to the attack rate discussed above: vaccination decreases the attack rate, leading to higher efficacy if the attack rate is high, and to lower efficacy if it is low.

#### S2.5 Sensitivity

To ensure the reliability and generality of our conclusions, we show the effect of changing values of several parameters. Namely, we consider: a different test sensitivity, by considering PCR instead of antigenic tests; an increase in the number of weekly introductions, from one every two weeks to one every week; vaccination coverage and a less effective vaccine; duration of the evaluation period.

##### S2.5.1 Sensitivity to test parameters

Here we consider a model for PCR tests. Symptomatic individuals remain isolated while they wait for their test results.

Table S5: Parameters for the considered interventions, with a PCR test [3]

| protocol parameter | value |
|---|---|
| $\theta_p$ | 0.8 |
| $\theta_c$ | 0.9 |
| $\theta_{sc}$ | 0.8 |
| $\Delta_Q$ | 7 days |
| $\Delta_R$ | 1 day |
| $\Delta_w$ | 1 day |

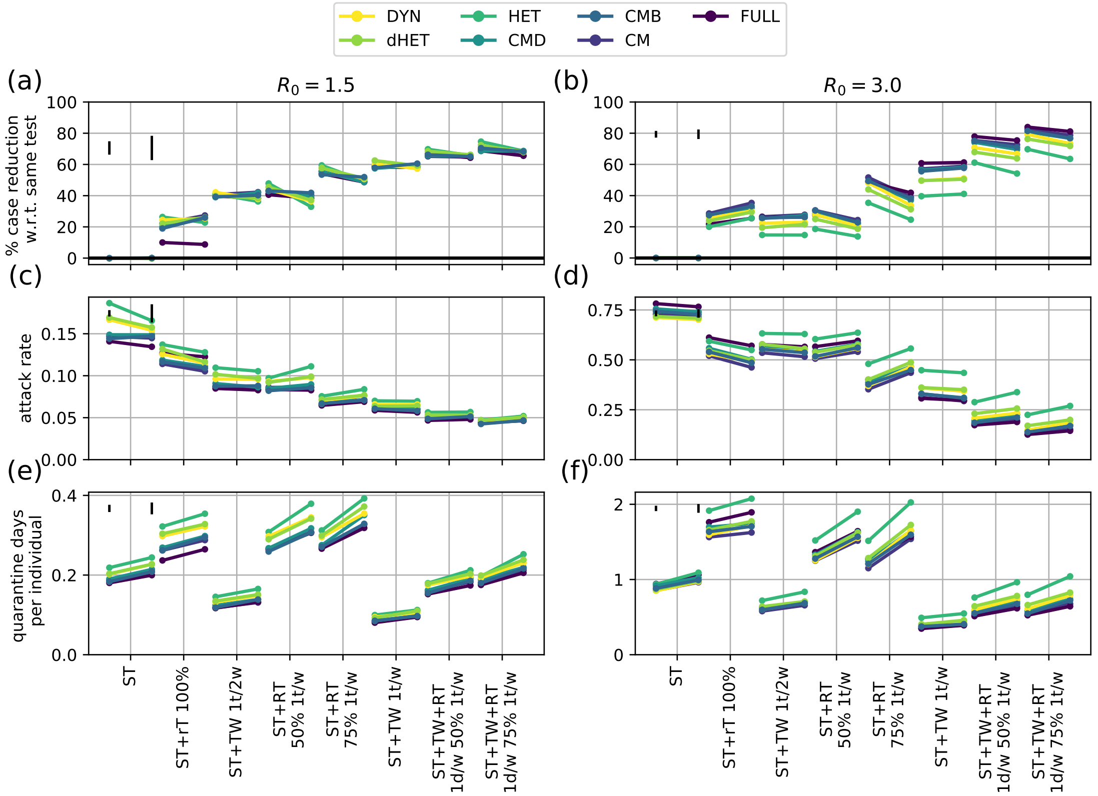

Figure S16: Sensitivity analysis on the impact of using a PCR test, for reproductive numbers $R_0 = 1.5$ (left) and $R_0 = 3$ (right), in the office setting. For each protocol, the left point correspond to values using antigenic tests, and the right one to PCR tests (parameters in Table S5). (a-b) Relative reduction with respect to ST with the same tests; (c-d) Attack rate at 60 days; (e-f) Days in quarantine per individual. Black vertical lines in the upper-left corner of each panel represent the median and maximum length of error bars among the data points.

##### S2.5.2 Sensitivity to the frequency of external introductions

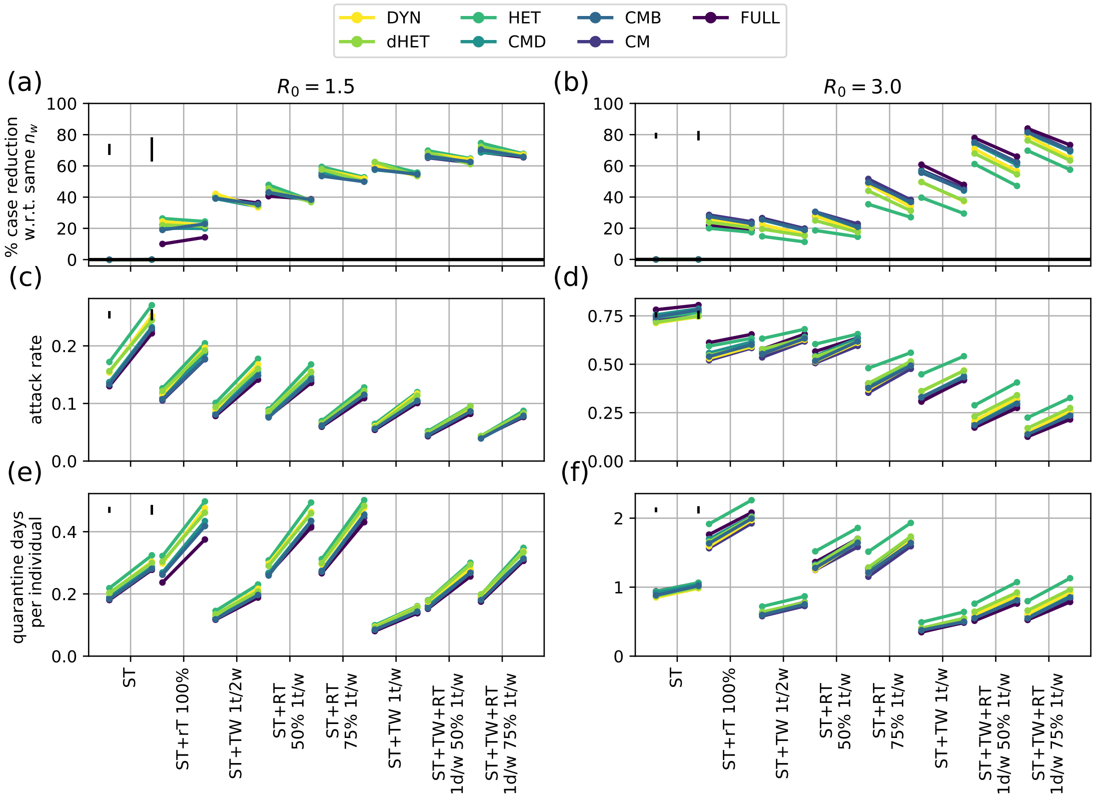

Figure S17: Sensitivity analysis on the impact of increased weekly introductions, for reproductive numbers $R_0 = 1.5$ (left) and $R_0 = 3$ (right), in the office setting. For each protocol, the left points corresponds to one introduction every two weeks, and the right point to one every week. (a-b) Relative reduction with respect to ST with the same frequency of introductions; (c-d) Attack rate at 60 days; (e-f) Days in quarantine per individual. Black vertical lines in the upper-left corner of each panel represent the median and maximum length of error bars among the data points.

##### S2.5.3 Sensitivity to vaccination coverage and a less effective vaccine

Table S6: Reduction in susceptibility $\sigma$, probability of clinical infection $p_c$ and relative infectiousness $r_\beta$, for a less effective vaccine [3]

| parameter | reduction |
|---|---|
| $\sigma$ | 85 % |
| $p_c$ | 93 % |
| $r_\beta$ | 50 % |

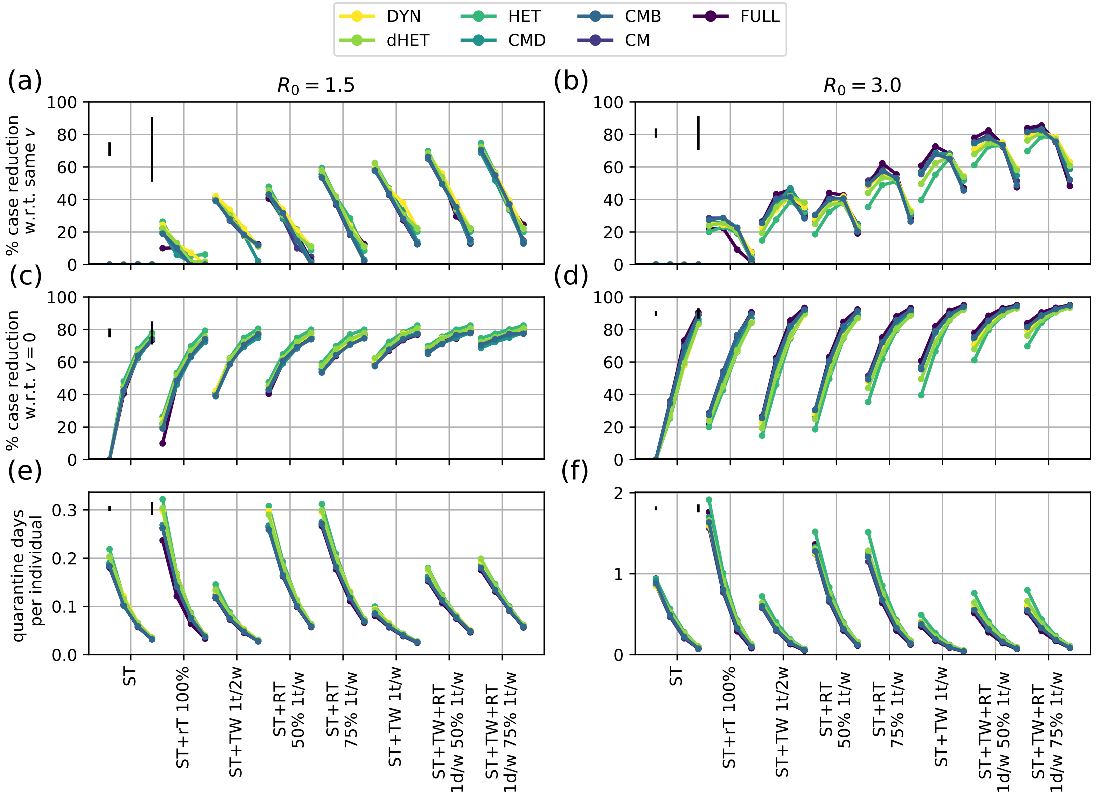

Figure S18: Effect of vaccine coverage for different NPIs in offices, using a less effective vaccine, for $R_0 = 1.5$ (left) and $R_0 = 3$ (right). For each protocol, points represent a coverage of 0%, 25%, 50%, and 75% from left to right. (a-b) Relative reduction with respect to ST with the same level of vaccine coverage; (c-d) Relative reduction with respect to ST with 0% vaccine coverage; (e-f) Days in quarantine per individual. Black vertical lines in the upper-left corner of each panel represent the median and maximum length of error bars among the data points.

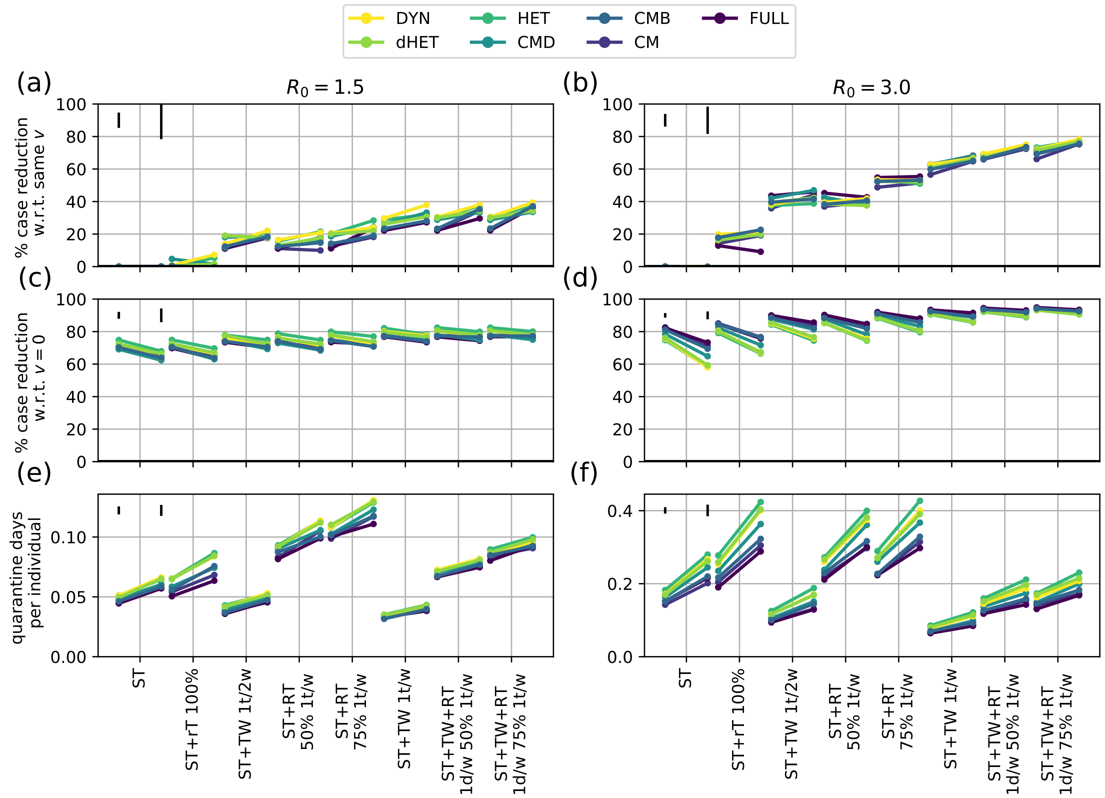

Figure S19: Comparing two type of vaccines at 50% coverage for different NPIs in offices, for $R_0 = 1.5$ (left) and $R_0 = 3$ (right). For each protocol, the left point corresponds to the vaccine considered in the main text, and the right point to a less effective vaccine (Table S6). (a-b) Relative reduction with respect to ST with the same level of coverage and type of vaccine; (c-d) Relative reduction with respect to ST with 0% vaccine coverage; (e-f) Days in quarantine per individual. Black vertical lines in the upper-left corner of each panel represent the median and maximum length of error bars among the data points.

##### S2.5.4 Sensitivity to the duration of the evaluation period

In the main text, we considered a duration of 60 days to evaluate NPIs. In figure S20, we show the effect of using different duration for the evaluation period, namely, 30, 60, 90 and 120 days. The value of the efficacy changes between 30 and 60 days, with an underestimation if measured only after 30 days, but remains stable when comparing 60, 90 and 120 days.

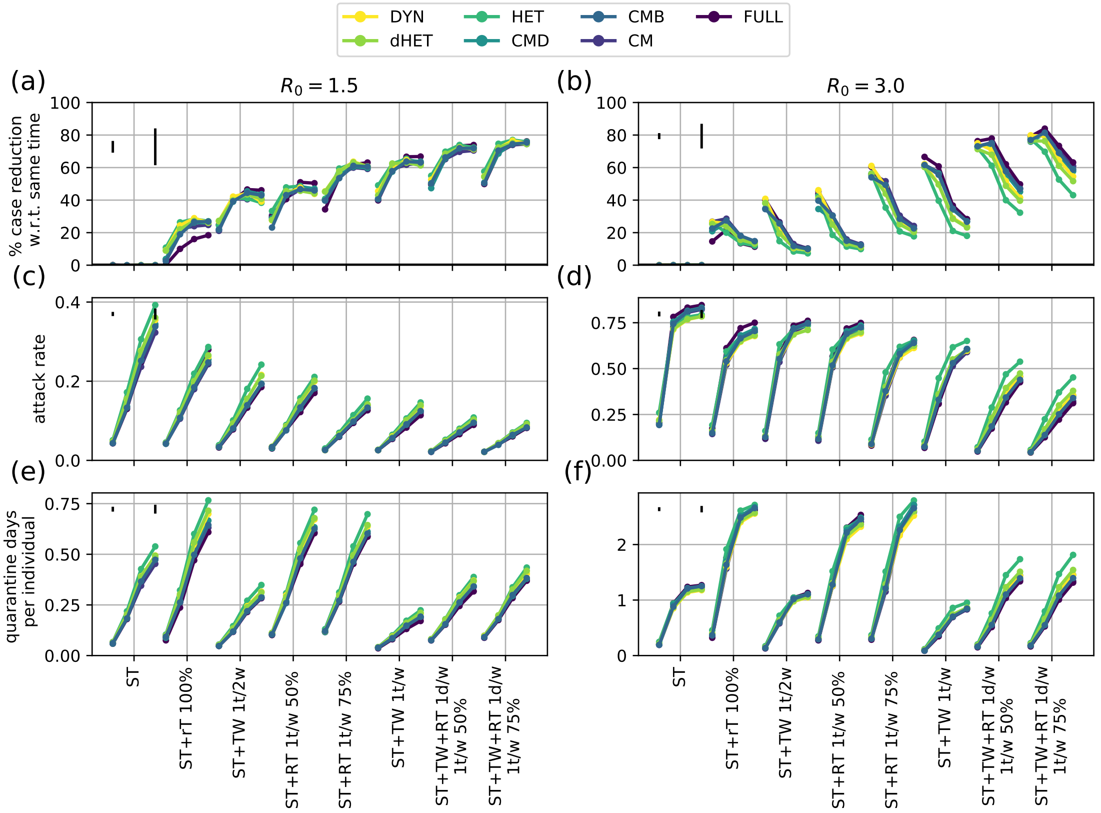

Figure S20: Comparing different time windows for NPI evaluation in offices, for $R_0 = 1.5$ (left) and $R_0 = 3$ (right). For each protocol, points correspond to a period of 30, 60, 90, and 120 days from left to right. (a-b) Relative reduction with respect to ST with the same parameters; (c-d) Attack rate; (e-f) Average number of days in quarantine per individual. Black vertical lines in the upper-left corner of each panel represent the median and maximum length of error bars among the data points.

### References

[1] Ciro Cattuto, Wouter Van den Broeck, Alain Barrat, Vittoria Colizza, Jean-François Pinton, and Alessandro Vespignani. Dynamics of person-to-person interactions from distributed rfid sensor networks. *PLoS ONE*, 5(7):e11596, 07 2010.

[2] Anna Machens, Francesco Gesualdo, Caterina Rizzo, Alberto E. Tozzi, Alain Barrat, and Ciro Cattuto. An infectious disease model on empirical networks of human contact: bridging the gap between dynamic network data and contact matrices. *BMC Infectious Diseases*, 13(1):1–18, 2013.

[3] Elisabetta Colosi, Giulia Bassignana, Diego Andrés Contreras, Canelle Poirier, Simon Cauchemez, Yazdan Yazdanpanah, Bruno Lina, Arnaud Fontanet, Alain Barrat, and Vittoria Colizza. Self-

testing and vaccination against covid-19 to minimize school closure. *Lancet Inf. Diseases*, in press, 2022.

[4] Anthony Christopher Davison and David Victor Hinkley. *Bootstrap methods and their application.* Number 1. Cambridge university press, 1997.

[5] Juliette Stehlé, Nicolas Voirin, Alain Barrat, Ciro Cattuto, Vittoria Colizza, Lorenzo Isella, Corinne Régis, Jean-Francois Pinton, Nagham Khanafer, Wouter Van den Broeck, and Philippe Vanhems. Simulation of an seir infectious disease model on the dynamic contact network of conference attendees. *BMC Medicine*, 9(1):87, 2011.
